# Device assessed 24-hour movement behaviour and cardiovascular disease mortality amongst cancer survivors

**DOI:** 10.64898/2026.06.09.26355299

**Authors:** John J Mitchell, Raaj Kishore Biswas, Joanna M Blodgett, Nicholas A. Koemel, Matthew N. Ahmadi, Abigail Fisher, Christine M. Friedenreich, Karen Canfell, I-Min Lee, Peter A. Cistulli, Dorothea Dumuid, Anthony D. Okely, Armando Teixeira-Pinto, Julia Steinberg, Anne E Cust, Emmanuel Stamatakis, Mark Hamer

## Abstract

**Background:** Cancer survivors face elevated risks of mortality from cardiovascular disease (CVD). The potential importance of physical activity (PA) and other behaviours across the 24-hour day (e.g. sedentary behaviour (SB) and sleep) for CVD-mortality risk is not well understood in this at-risk population.

**Objectives:** To assess the importance of 24-hour movement behaviour, using a compositional approach, for mitigating CVD-mortality amongst cancer survivors.

**Methods:** Participants with a prior cancer diagnosis were drawn from the UK Biobank accelerometry sub-study (n=6,158). Accelerometer-derived movement (moderate-to-vigorous PA (MVPA), vigorous PA (VPA), moderate PA (MPA), light PA (LPA), SB, sleep) was examined in relation to CVD-mortality, identified from health record linkage data (using Fine-Gray Cox proportional-hazards models adjusted for demographic, health, lifestyle covariates).

**Results:** Median follow-up was 8.0 years (Q1-Q3: 7.4-8.5), with n=500 (8.2%) deaths (CVD-deaths: n=118). Greater MVPA, in place of any other behaviour, was inversely associated with CVD-mortality with e.g. 10% lower hazard if MVPA theoretically replaced 7 minutes (mins)/day SB (Hazard ratio (HR): 0.91, (95% Confidence Interval: 0.86-0.95)), 9 mins/day LPA (HR: 0.90, 0.83-0.97), or 11 mins/day sleep (HR: 0.90, 0.83-0.97). The VPA component of MVPA proved critical, requiring only ∼1-2 additional mins/day for equivalent hazard reduction. Sleep duration, was also inversely associated with CVD-mortality. A 10% lower hazard required replacing 29 mins/day of SB with sleep (HR: 0.90, 0.84-0.96); no other behavioural replacement amongst SB, sleep or LPA could provide an equivalent risk reduction.

**Conclusions:** Among cancer survivors, the most potent reduction in CVD-mortality followed theoretically reallocating time to higher intensity movement.

## Introduction

Cancer survivorship is increasing globally because of advances in early detection and improved treatments^1,2^. However, survivors face elevated risks of both cancer-related and non-cancer-related morbidity and mortality, particularly cardiovascular disease (CVD) and other metabolic disorders^3,4^. It is estimated that more than 50% of survivors will develop at least one other comorbidity within 5-years post-diagnosis^5^. The importance of physical activity (PA) for CVD in the general population is established^6^, with an increasing recognition of the broader importance of all 24-hour movement behaviours, including minimising sedentary behaviour (SB) and increasing sleep^7–9^. Exercise training regimens have garnered attention as possible primary and secondary prevention strategies for subsequent CVD prevention in cancer survivors^10^. However, the role of PA within the wider context of all movement behaviour in the 24-hour day remains poorly understood.

A meta-analysis of 136 studies focussing on cancer survivors found higher levels of post-diagnosis PA to be inversely associated with subsequent cancer-specific mortality with some evidence of a dose-response association^11^, although CVD endpoints were less well-explored. One large-scale study has revealed the reduced risk of subsequent myocardial infarction and heart failure from cancer survivors who adopted PA after diagnosis^12^. However, the extant literature almost exclusively relies on self-reported measures of PA, which are prone to bias, and does not account for the wider elements of daily movement (e.g. sitting, sleep, lighter intensity PA) beyond leisure-time or exercise-specific PA volume^13–15^. For instance, SB has been associated with an increase in mortality risk amongst cancer survivors, independently of PA^15–17^. Sleep, despite often being disrupted amongst cancer survivors due to the complex biological disease status and treatment strategies^18^, has not been well-studied in relation to subsequent CVD-mortality. Both challenges can now be overcome using accelerometry, which accurately captures all daily movement behaviour in the 24-hour day. Despite this advancement, and the known interrelatedness of movement behaviours (e.g. greater active time must replace inactive time), and resulting competing advantages/disadvantages from different behaviours, the 24-hour movement paradigm has received limited interest as a tool for improving long-term survivorship^19^.

Evidence in large UK and US cohorts provides an insight to the 24-hour movement profile of cancer survivors by comparing them to cohort members with no history of cancer. These studies reveal major differences, including lower daily moderate-to-vigorous PA (MVPA) and light-intensity PA (LPA), but also greater SB and longer sleep duration in this subset^20–23^.

Despite the known differences, and evidence for independent effects of SB and PA on CVD-mortality in cancer survivors, studies have: *(i)* relied on low-resolution measures of PA and/or SB (e.g. self-reported, or not also separating vigorous PA (VPA) from moderate PA (MPA)) and, *(ii)* do not account for the wider impact from other movement behaviours of the 24-hour day, including sleep, and *(iii)* do not consider their differential effects on CVD-specific mortality, compared to all-cause or cancer-mortality. The primary aim of this study was to investigate associations between accelerometer-derived 24-hour daily movement behaviour and risk of CVD-specific mortality amongst cancer survivors, with secondary aims to examine whether associations differ across the intensity-gradient (e.g. MPA versus VPA), and finally to investigate these same associations with all-cause and cancer-mortality.

## Methods

### Study Sample

The sample of cancer survivors was drawn from UK Biobank (UKBB), a large prospective cohort comprising 502,629 individuals across the UK, aged 40–69 years at enrolment between 2006 and 2010^24^. Study inclusion was further restricted to participants from the UKBB accelerometry sub-study^25^, which received ethical approval from the UK National Research Ethics Service (No. 11/NW/0382) and recruited 103,684 individuals from June 2013 to 2015 to wear a wrist accelerometer device. All participants gave informed consent prior to completing health and lifestyle questionnaires and undergoing a series of physical assessments conducted by trained staff. Participants were included if they had any history of cancer, occurring prior to their accelerometer collection. Incidence and date of diagnosis was derived via linkage with the National Health Service (NHS) Digital records for participants based in England and Wales, while the NHS Central Register was used to identify cancer events in participants in Scotland. Inpatient hospitalisation data were available from the Hospital Episode Statistics for England, Patient Episode Database for Wales, or the Scottish Morbidity Record for Scotland. For England and Wales, cancer diagnosis data were followed up to 31 December 2020 and 31 December 2016 respectively^24^, and for Scotland, diagnosis data were followed-up through 30 November 2021. To identify cancer survivors, health record linkage data were censored at the participant date of accelerometer-wear. Where linkage was not available, this was supplemented with self-reported cancer history (n=2,185). ICD-10 codes and site-specific cancer events from linked data (n=3,973) within the accelerometry sub-study are presented in table S1. Sample derivation is presented in figure S1.

### Measurement of 24-hour movement behaviour

Movement behaviour was collected using an Axivity AX3 accelerometer device (Axivity, Newcastle, UK) worn on participant’s dominant wrist for up to 7 days^25^ with a sampling frequency of 100Hz. A minimum of 16 hours of wear time was considered a valid wear-day in alignment with previous studies in UKBB^26^, and participants were included if they wore the device for a minimum of 3 wear-days including a weekend day^27^. Initial processing involved correction for orientation and removal of non-wear using standard procedures^27,28^. Waking time was differentiated from sleep periods using a sleep-algorithm, previously validated using sleep diaries and polysomnography^29^.

Physical activity intensity was classified with a two-level Random Forest algorithm^26,30^ (further details in supplementary text S1). The first level classifies posture type (moving, standing/standing semi-stationary and sedentary postures) in contiguous 10s bins^31,32^. The second level categorizes standing and moving activities into intensity bands. Moving and standing activities are then grouped by their acceleration into LPA, MPA, VPA, or MVPA^26^.

### Compositions of 24-hour daily movement behaviour

Two compositions of participant daily movement behaviours were derived reflecting average daily time in minutes spent in each behaviour and normalised to 24 hours^9^ given wear-adherence was high (average daily wear time: 23h, 43m). The first composition was a 4-part composition encompassing MVPA, LPA, SB and sleep time. Given findings that VPA accrued in short bouts under 1 minute long have shown strong associations with incident cancer^26^ and CVD^33^, and the interest in purposeful exercise interventions^10^, vigorous PA (VPA) and moderate PA (MPA)^32^ were disaggregated in a second, nested, five-part composition of activity (VPA, MPA, LPA, SB, sleep), providing a better understanding of the differential associations across the PA intensity gradient.

### Outcome

The primary outcome was CVD-specific mortality and secondary outcomes were all-cause and cancer-specific mortality, identified through linkage with participant health records (Cardiovascular ICD-10 codes: I00–I99, I11, I13, I20-I51, I60-I69; Cancer CD-10 codes: C00–C97; Table S1). Participants were followed up through to November 30th, 2022.

### Statistical Analysis

Comparisons of the compositions between cancer survivors with a mortality event and those without, are displayed using log-ratio difference plots with bootstrapped 95% confidence intervals (B=1000 bootstraps). An established isometric log-ratio (ILR)-pivoting approach^9,34^ (described further in supplementary text S2) with a Cox-proportional hazards approach was used to model the associations between 24-hour movement compositions and CVD-mortality, all-cause mortality and cancer-mortality separately. Examination of cause-specific mortality additionally included a Fine-Gray sub-distribution of hazards approach to account for competing risks. Two models were fitted, including: (a) age and sex-adjusted, and (b) after additionally adjusting for demographic, health and lifestyle covariates: ethnicity, highest attained level of education, family history of cancer, medication use, previous history of CVD, smoking status, diet quality and alcohol use (Table S2). Only the fully-adjusted model is reported in-text. BMI was excluded as a covariate given its likely position on the causal pathway between PA volume and long-term adverse health outcomes^35^.

Isotemporal substitutions aid in interpretation of compositional models by estimating the change in mortality hazard after minute-by-minute reallocation of time away from or into the significant behaviour, while holding all other behaviours fixed at the sample average. It is only presented for behaviours that were statistically significant, relative to all other behaviours, in fully-adjusted models. To provide meaningful examples the time-reallocations associated with 10 and 20% reduction/increase in Hazard Ratio (HR) are reported in-text.

### Sensitivity analyses

To mitigate for possible reverse causality, we removed all individuals with only self-reported cancer events, any deaths occurring in the first two years of follow up^35^ as well as any individuals with any cancer diagnosis that occurred during the two years preceding baseline accelerometry measurement. Secondly, individuals with non-melanoma skin cancers were removed. Finally, the degree of residual confounding was assessed by calculating E-values, which estimate the strength of association necessary between an unmeasured confounder and both the exposure and outcome to nullify any significant associations between the lead ILR-coordinate and the outcome^36^.

## Results

### Participant demographics

A total of 6,158 individuals with a previous history of any cancer prior to wearing the accelerometer device were included (Figure S1). The median age at baseline was 64.9 years (SD: 7.0) and the median follow-up was 8.0 years (Q1-Q3: 7.4-8.5).

Median time from primary cancer to follow-up was 6.97 years (Q1-Q3: 2.8-10.7). Participants were predominantly female (66%), a large proportion held a college or university degree (42%) and a similar proportion had never smoked, (40%) and reported alcohol consumption within guideline levels (59%) (Table 1). At baseline, 29% of individuals reported taking antihypertensives, anti-hypercholesterolaemia or insulin and 13% had a documented history of CVD. The most commonly diagnosed cancer sites among survivors were breast (n = 1298; 33%), non-melanoma skin (n = 674; 17%), and prostate cancer (n = 410; 10%). During the follow-up window, 500 participants (8.2%) subsequently died from any cause, which included 118 (1.9%) deaths from cardiovascular disease and 375 (6.1%) deaths from cancer.

**Table 1.** Baseline characteristics of included UK Biobank participants at accelerometry assessment (n = 6,158), stratified by mortality outcomes (cardiovascular, all-cause and cancer)

|  |  |  |  |  |  |  |  |  |  |
| --- | --- | --- | --- | --- | --- | --- | --- | --- | --- |
|  |  | **Total Sample**<br>n=6,158 |  | **CVD-Mortality**<br>n=118 |  | **All-cause Mortality**<br>n=500 |  | **Cancer-Mortality**<br>n=375 |  |
| **Age (years)** | Mean (SD) | 64.9 | (7.0) | 68.5 | (6.0) | 67.5 | (6.2) | 67.1 | (6.2) |
| **Sex (female)** | n % | 4074 | 66% | 46 | 39% | 228 | 46% | 184 | 49% |
| **Ethnicity (White)** | n % | 6012 | 98% | 113 | 96% | 479 | 96% | 362 | 97% |
| **Educational attainment** | n % |  |  |  |  |  |  |  |  |
| College/University degree |  | 2576 | 42% | 38 | 32% | 192 | 38% | 157 | 42% |
| A/AS-level |  | 736 | 12% | 17 | 14% | 63 | 13% | 44 | 12% |
| NVQ/HND/HNC |  | 302 | 5% | 10 | 8.50% | 35 | 7% | 20 | 5% |
| CSE |  | 189 | 3% | 5 | 4.20% | 13 | 3% | 7 | 2% |
| O-levels |  | 1351 | 22% | 25 | 21% | 104 | 21% | 78 | 21% |
| Other education |  | 1004 | 16% | 23 | 19% | 93 | 19% | 69 | 18% |
| **Smoking** | n % |  |  |  |  |  |  |  |  |
| Current |  | 390 | 6.30% | 12 | 10% | 45 | 9% | 29 | 7.70% |
| Never |  | 2459 | 40% | 59 | 50% | 231 | 46% | 169 | 45% |
| Ex |  | 3309 | 54% | 47 | 40% | 224 | 45% | 177 | 47% |
| **Medication use** | n % | 1777 | 29% | 64 | 54% | 222 | 44% | 157 | 42% |
| **Alcohol Consumption** | n % |  |  |  |  |  |  |  |  |
| Above guidelines |  | 2179 | 35% | 42 | 36% | 197 | 39% | 146 | 39% |
| Ex |  | 179 | 2.90% | 5 | 4.20% | 22 | 4.40% | 17 | 4.50% |
| Never |  | 3611 | 59% | 64 | 54% | 261 | 52% | 200 | 53% |
| Within guidelines |  | 189 | 3.10% | 7 | 5.90% | 20 | 4% | 12 | 3.20% |
| **Family history of cancer** | n % | 4384 | 71% | 89 | 75% | 360 | 72% | 261 | 70% |
| **Diet Quality** | Median (Q1-Q3) | 8 | (6–10) | 7 | (5–10) | 7 | (5–10) | 7 | (5–10) |
| **Previous CVD** | n % | 771 | 13% | 50 | 42% | 135 | 27% | 94 | 25% |
| Abbreviations. CVD: Cardiovascular disease; PA: Physical activity Q: Quartile. |  |  |  |  |  |  |  |  |  |

### Participant 24-hour movement profiles at baseline

The average 24-hour movement composition of the cohort was 27 minutes (mins) of MVPA, 4 hours (h) 57mins of LPA, 10h 53mins of SB, and 7h 43m of sleep (Table S3). Across all mortality outcomes, individuals who died during follow-up had, on average, accumulated less MVPA and LPA and more SB than survivors (Figure 1). For example, compared with the average composition of survivors, those who subsequently died from CVD had accrued 53% less MVPA (95% Confidence Interval (CI): -69%, -36%; ∼15 fewer mins/day), 7% less LPA (-11%, -3%; ∼20 fewer mins/day), 8% more SB (6%, 11%; ∼55 additional mins/day), and 6% less sleep (-9%, -2%; ∼25 fewer mins/day). Differences were directionally similar but smaller for all-cause mortality and cancer-mortality.

**Figure 1.**
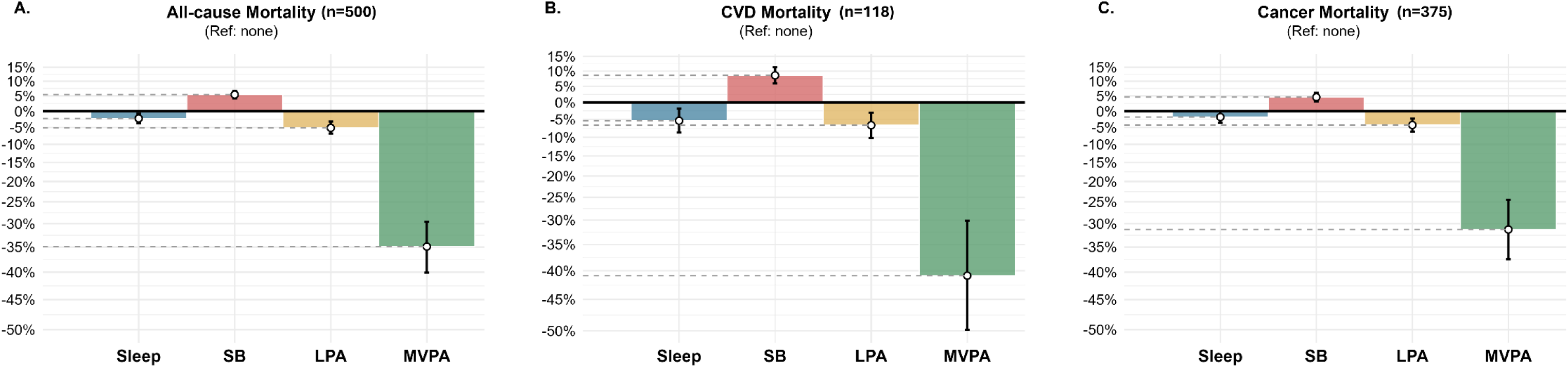
Relative differences in 24-hour time use by mortality status (compared to cancer survivors with no mortality event). ***Legend:*** Proportional differences in 24-hour movement compositions among cancer survivors (n=6,158) with bootstrapped 95% confidence intervals. A: all-cause mortality (n=500); B: CVD-mortality (n=118); C: Cancer-mortality (n=375). Median follow-up 7.97 years (Q1-Q3: 7.4-8.5).

When MVPA was disagregated to MPA and VPA, participants on average spent 2 daily mins in VPA and 25 daily mins in MPA (Table S4, Figure S2). On average, individuals who subsequently died of CVD, accrued 53% less VPA (-41%, -63%; 1-2 fewer mins/day) and 40% less MPA (-29, -49%; ∼10 fewer mins/day). Differences were similar for all-cause mortality and cancer-mortality (Figures 1,S2).

### Associations of 24-hour movement profiles with mortality

Greater time spent in MVPA, relative to all other movement behaviours, was associated with lower CVD-mortality risk. Greater SB, relative to all other behaviours, was associated with higher CVD-mortality (Tables S5-S7). LPA, relative to all other behaviours, was not associated with any outcome. Longer sleep duration, relative to all other behaviours, was inversely associated with CVD-mortality. Similar associations were observed for all-cause and cancer-specific mortality outcomes among cancer survivors, but associations for sleep duration proved null for these secondary outcomes.

The difference in HR associated with reallocating time into and out of MVPA from the sample daily average was asymmetric in magnitude. For example, a ∼10% lower hazard of CVD-mortality was observed after MVPA replaced 7 mins of SB (HR:0.91, 95%CI: 0.86-0.95, Figure 2, Table S8), 9 mins of LPA (HR:0.90, 0.83-0.97), or 11 mins of sleep (HR:0.90, 0.83-0.97). A ∼20% lower hazard was observed after MVPA replaced 18 mins of SB (HR: 0.80, 95%CI: 0.72-0.89), 22 mins of LPA (HR:0.80, 0.67-0.95), or 30 mins of sleep (HR: 0.80, 0.75-0.85). Conversely, a ∼10% greater hazard was observed when MVPA was replaced by 6 daily mins of SB (HR: 1.11, 1.05-1.17) or LPA (HR:1.10, 1.03-1.17), or 7 mins of sleep (HR:1.10, 1.03-1.18). A ∼20% greater hazard was observed after MVPA was reduced by 10 mins of SB (HR: 1.21, 1.10-1.33) or LPA (HR: 1.19, 1.06-1.33), or 11 mins of sleep (HR: 1.19, 1.06-1.33).

**Figure 2.**
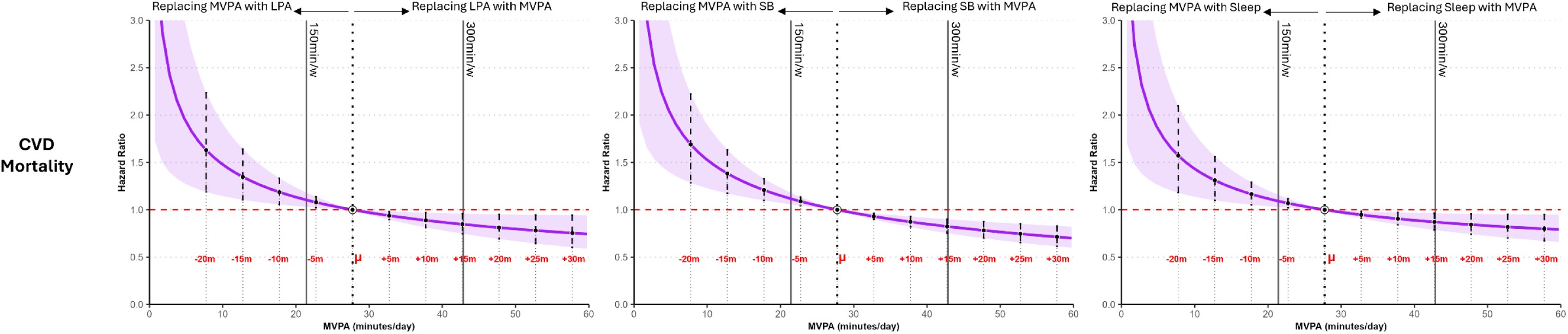
Difference in hazard of CVD-mortality from theoretical reallocation of time into/out of MVPA amongst cancer survivors. ***Legend:*** Hazard ratios of CVD-mortality from time reallocations between MVPA and each of LPA, SB, and sleep among n=6,158 cancer survivors centred at the sample mean daily MVPA (white dot; µ). Fine–Gray models were adjusted for age, sex, ethnicity, smoking, alcohol use, diet quality, family history of cancer, CVD history, and regular medication use.

Similar estimated effects from time reallocation were observed for all-cause and cancer-mortality, despite larger reallocations needed to demonstrate comparable risk reductions (Tables S9,S10 & figures S3,S4).

Beyond MVPA, the only other behavioural change associated with a lower CVD-mortality risk was reallocating SB to sleep (Figure 3). A ∼10% lower hazard was observed after reallocating 67 mins/day of SB into sleep (HR: 0.90, 0.83-0.98, Table S10), and 20% observed after a total of 2h 19 mins/day (HR: 0.80, 0.67-0.95).This association was weaker in magnitude for all-cause mortality (Table S12) and null for cancer-mortality.

**Figure 3.**
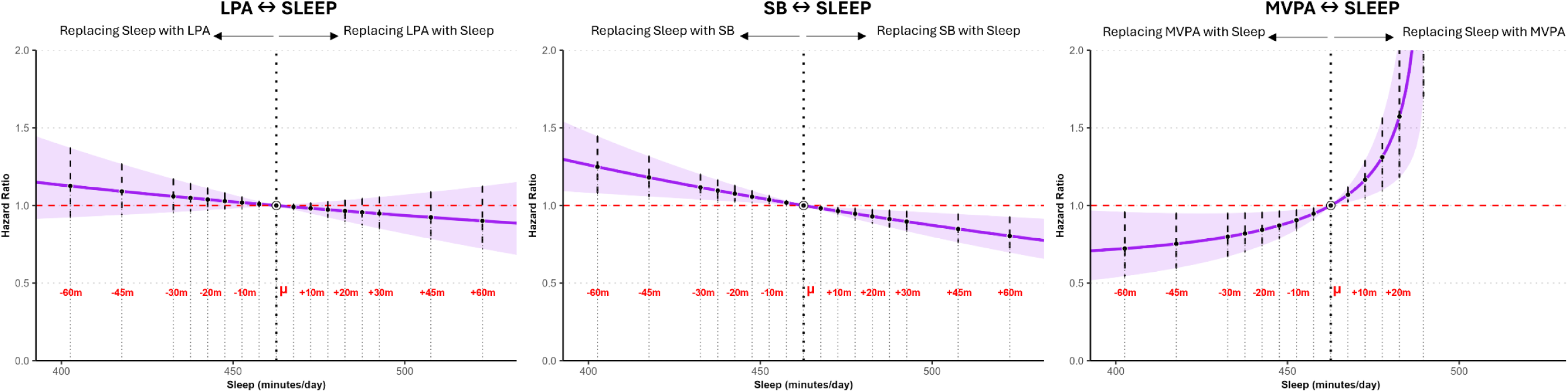
Difference in hazard of CVD-mortality from theoretical reallocation of time into/out of sleep amongst cancer survivors. ***Legend:*** Hazard ratios for CVD-mortality from time reallocations between sleep and each of LPA, SB, and MVPA among n = 6,158 cancer survivors, centred at the sample mean daily sleep (white dot; µ). Fine–Gray models were adjusted for age, sex, ethnicity, smoking, alcohol use, diet quality, family history of cancer, CVD history, and regular medication use.

### Differential associations across the physical activity intensity gradient

When data were re-analysed with MVPA disaggregated into VPA and MPA, only VPA (relative to the remaining behaviours) was inversely associated with CVD-mortality, while both VPA and MPA (each relative to the remaining behaviours) were inversely associated with all-cause mortality and cancer-mortality (Tables S13-S15).

The change in HR associated with reallocating time into VPA was steep at low levels of activity. For example, a ∼10% lower hazard of CVD-mortality was observed after VPA increased by 1 min/day in place of any behaviour, including MPA (HR: 0.92, 95%CI: 0.86-0.98; Figure S5, Tables S16). A ∼20% lower hazard was observed after VPA increased by 3 mins/day when replacing SB (HR: 0.81, 95%CI: 0.70-0.94), or by 4 mins/day when replacing MPA (HR: 0.80, 0.65-0.99), LPA (HR: 0.79, 0.66-0.94), or sleep (HR: 0.79, 0.66-0.95). Replacement of 2 mins/day of VPA with any behaviour including MPA (constituting loss of all average daily VPA), was associated with >20% greater hazard of CVD-mortality (HR: 1.22, 1.00-1.48).

Similar estimated effects of VPA from time reallocation were observed for all-cause and cancer-mortality (Tables S17-18). Alternatively for both outcomes, greater MPA in place of any behaviours other than VPA required much greater (8-13 mins) time replacement to observe even 10% lower hazard (Figure S6, Table S19-20).

### Sensitivity analysis

After exclusion of all self-reported cancers (n=2185) individuals with any cancer episode occurring within 2y prior to accelerometry (n=481) or any deaths occurring in the first 2y of follow up (n=27 cancer-mortality events), which may reflect reverse causation, showed subtle differences. Sleep relative to other behaviours now spanned the null. For cancer-mortality only, SB, relative to all other behaviours, was no longer associated with cancer-mortality (Tables S21-S23). No findings were substantively changed by removal of participant’s with a primary event of non-melanoma skin cancer (n=674) (Tables S24-26). E-values indicated that a residual confounder would need to have moderate associations with both the exposure and outcome to otherwise explain the reported associations. For MVPA, E-values ranged from 1.73 to 1.83 across outcomes. E-values were larger for SB and sleep, particularly for CVD-mortality (E-value = 3.14 and 2.45, rspectively), suggesting robustness to unmeasured confounding (Tables S27-29).

## Discussion

This study examined associations between 24-hour movement behaviours and subsequent CVD-mortality, cancer-mortality and all-cause mortality amongst cancer survivors. Individuals who died during follow-up were less active at baseline, in particular accumulating substantially lower (40–50%) MVPA, and greater (5–8%) SB, relative to cancer survivors with no subsequent mortality event. This pattern was most prominent amongst cancer survivors with a subsquent CVD-mortality event compared to those who died of other causes. Greater MVPA, relative to all other behaviours in the 24-hour day, was inversely associated with subsequent CVD-mortality. Even small increases in MVPA (7–8 mins/day) in place of LPA, SB, or sleep corresponded to ∼10% lower hazard of CVD-mortality. However, this association appeared underpinned by the VPA component (over MPA), with only 1-2 additional mins/day VPA being associated with this same reduction in hazard. The inverse pattern was similarly severe; losing as little as 5–9 mins/day of MVPA (or 1-2 minutes of daily VPA) was associated with a 10–20% higher CVD-mortality hazard, underscoring the disproportionate importance of attaining MVPA, and particularly VPA in this population. The only other time-exchange associated with comparable reduction in CVD-mortality risk involved replacing substantive (>60 minute) amounts of SB with sleep. Together, these findings suggest that preserving even small amounts of daily MVPA may be critical for subsequent longevity among cancer survivors, but particularly the VPA component for mitigating subsequent risk of CVD-mortality.

The observed baseline compositional differences in 24-hour movement profiles align with previous findings within UKBB^20^, and our findings extend this research by examining their relationship with subsequent mortality within cancer survivors. Our findings align with and build on previous meta-analyses^37,38^ and trials^38,39^ by addressing a key methodological gap, and focussing on CVD-mortality which has received much less attention but may have physiologically unique pathways from other causes of subsequent mortality. While previous evidence has been used to recommend PA in formal guidelines for cancer survivors^40,41^ those guidelines were based mainly on research studies that used questionnaire-derived PA, and none had examined the full spectrum of daily movement, including sleep duration^38,42–44^. This study meanwhile utilises the highest resolution movement data yet available to investigate possible alternative pathways (e.g. involving SB and sleep) to enhance postdiagnostic survival. The findings (i) reinforce the importance of MVPA for reducing subsequent mortality in this at-risk group, (ii) reveal replacing sedentary time with sleep to be a novel, alternative potential behavioural pathway for longevity in cancer survivors worthy of investigation, and (iii) identify VPA as the critical intensity, over MPA or LPA, for reducing risk of subsequent CVD-mortality.

Many of the hypothesized biologic pathways between daily movement behaviours, sleep and subsequent mortality in cancer survivors are similar to those that exist for the general population. These mechanisms include global improvements in cardiometabolic health and reduced systemic inflammation^9,45^. However, cancer survivors represent a higher-risk group, with greater cardiometabolic vulnerability often arising from treatment-related cardiotoxicity, accelerated vascular ageing, inflammation, and immune dysregulation^46,47^, which may explain the large observed effect sizes for CVD-mortality, principally driven by VPA. These mechanisms are critical for cardio-oncology survivorship care, suggesting that higher intensity PA may be required to meaningfully restore and maintain cardiorespiratory fitness and vascular function among cancer survivors^9^. This finding aligns with the observed cardio-protective effects of even minimal VPA accrued incidentally (<1 minute bouts) in the general population^48^. These findings reinforce the importance of MVPA for influencing survivorship outcomes over and above those that are operative in the general population, but highlight VPA as particulary critical for post-diagnostic cardiovascular health; a major challenge for the field given the practical difficulties facing many cancer survivors wishing to engage in more vigorous activities.

Finally, this study may implicate sleep as a potentially important and distinct behavioural target among cancer survivors. Although survivors may on average have longer sleep durations^20^, growing evidence indicates that sleep quality, including regularity (unable to be assessed in the present study) is frequently impaired^49^ and is important for health and longevity^50^. Survivors report a higher prevalence of fragmentation, insomnia symptoms, and non-restorative sleep compared with the general population^49^. Sleep dysregulation has been linked to many of the same cardiometabolic pathways mentioned above including systemic inflammation, and elevated blood pressure^51^. While reallocating SB to additional sleep may simply help to reduce the cardiometabolic harms of sitting, we hypothesise that longer sleep durations may also compensate for qualitative sleep deficits. Further large-scale prospective studies examining both duration and quality in cancer survivors are key to understanding this association.

### Strengths/limitations

This study has a number of strengths including the large sample size of the UKBB and the use of validated accelerometry algorithms with high classification precision^30^ which permit investigation into daily movement behaviour in an unprecedented level of granularity. Further, this study used a gold-standard compositional analyses^34^. The extended follow-up period within UKBB allowed us to address many of the methodological limitations of prior literature including a robust examination of reverse causation.

Limitations include the recognised sample selection bias observed in UKBB^52^. The possibility of residual confounding also cannot be excluded. Most notably, the lack of data regarding cancer treatment, with known cardiotoxic effects of many anti-cancer therapies^46^. Further, this study necessarily relies on a one-week accelerometry measurement period, and there is inevitable possibility for behaviour misclassification, despite the high specificity of movement classification algorithms^30^.

### Conclusions

Recent cardio-oncology frameworks emphasise the importance of exercise as a strategy to prevent and manage treatment-related cardiovascular toxicity^53^, yet existing evidence has largely focused on structured exercise interventions, or examined only leisure-time or MVPA^11,12^, rather than the broader distribution of daily movement behaviours. Our findings extend current evidence, revealing how even minimal changes in daily MVPA (and critically VPA) accrued within the 24-hour day are associated with reduced future CVD-mortality risk. Where increases in PA are not feasible, reducing prolonged sedentary time in favour of sleep may be a pragmatic adjunct strategy.

## Data Availability

The data that support the findings of this study are available from the UK Biobank, but restrictions apply to the availability of these data, which were used under license for the current study, and so are not publicly available.

https://www.ukbiobank.ac.uk/use-our-data/apply-for-access/

## Declarations

### Author Contributions

JJM, RB, and MA had full access to the data and conducted the analyses. Study concept and design were developed by JJM, ES, MA, NK, and MH. Data acquisition, analysis, or interpretation was undertaken by JJM, RB, MA, and JMB. The manuscript was drafted by JJM. All authors contributed to critical revision of the manuscript for important intellectual content. Statistical analyses were performed by JJM, RB, NK, and MA. Funding was obtained by ES and MH. Administrative, technical, or material support was provided by ES and MA. Study supervision was provided by ES and MH.

## Supplementary materials

Supplementary material_V8.docx

