## Supplement for "Device assessed 24-hour movement behaviour and cardiovascular disease mortality amongst cancer survivors"

Contents

Table S1a Cancer history ICD-10 codes. 4

Table S1b. Cancer sites of cancer survivor sample (excl. self-reported cases (n=2185)). 5

Supplementary text S1. Sleep and Physical activity intensity classification algorithms. 6

Supplementary text S2. Compositional 'pivot' approach 7

Table S2 Included Covariate definitions. 8

Figure S1 Sample derivation for UK Biobank participant inclusion resulting in n=6,158 included participants. 9

Table S3. Movement behaviour, and difference from the sample compositional average, by outcome. 10

Table S4. Movement behaviour (separated by intensity), and difference from the sample compositional average, by outcome. 11

Figure S2. Relative differences in 24-hour time use (separated by intensity) by mortality status (compared to cancer survivors with no mortality event). 12

Table S5. Association of Coordinates of daily-movement behaviours (intensities, including MVPA) and sleep with risk of CVD-mortality among previous cancer survivors (Fine-Gray Cox Proportional Hazards Model) 13

Table S6. Association of Coordinates of daily-movement behaviours (intensities, including MVPA) and sleep with risk of All-cause mortality among previous cancer survivors (Cox Proportional Hazards Model) 14

Table S7. Association of Coordinates of daily-movement behaviours (intensities, including MVPA) and sleep with risk of cancer-mortality among previous cancer survivors (Fine-Gray Cox Proportional Hazards Model) 15

Table S8. Predicted HR of CVD-mortality amongst Cancer survivors from Isotemporal substitution around sample average of MVPA whilst holding other components constant at sample average. 16

Figure S3. Difference in hazard of subsequent All-cause mortality from theoretical reallocation of time into/out of MVPA amongst cancer survivors. 17

Table S9. Predicted HR of All-cause mortality amongst Cancer survivors from Isotemporal substitution around sample average of MVPA whilst holding other components constant at sample average. 17

Figure S4. Difference in hazard of subsequent cancer-mortality from theoretical reallocation of time into/out of MVPA amongst cancer survivors. 18

Table S10. Predicted HR of Cancer-specific mortality amongst former Cancer survivors from Isotemporal substitution around sample average of MVPA whilst holding other components constant at sample average. 18

Table S11. Predicted HR of CVD-mortality amongst Cancer survivors from Isotemporal substitution around sample average of Sleep whilst holding other components constant at sample average. 20

Table S12. Predicted HR of All-cause mortality amongst Cancer survivors from Isotemporal substitution around sample average of Sleep whilst holding other components constant at sample average. 21

Table S13. Association of Coordinates of daily-movement behaviours (intensities) and sleep with risk of CVD-mortality (Fine-Gray Cox Proportional Hazards Model) 22

Table S14. Association of Coordinates of daily-movement behaviours (intensities) and sleep with risk of All-cause mortality (Fine-Gray Cox Proportional Hazards Model) 23

Table S15. Association of Coordinates of daily-movement behaviours (intensities) and sleep with risk of cancer-mortality (Fine-Gray Cox Proportional Hazards Model) 24

Figure S5. Difference in hazard of all mortality outcomes from theoretical reallocation of time into/out of VPA amongst cancer survivors. 25

Table S16. Predicted HR of CVD-mortality amongst Cancer survivors from Isotemporal substitution around sample average of VPA whilst holding other components constant at sample average. 26

Table S17. Predicted HR of All-cause mortality amongst Cancer survivors from Isotemporal substitution around sample average of VPA whilst holding other components constant at sample average. 27

Table S18. Predicted HR of subsequent cancer mortality amongst Cancer survivors from Isotemporal substitution around sample average of VPA whilst holding other components constant at sample average. 28

Figure S6. Difference in hazard of all mortality outcomes from theoretical reallocation of time into/out of MPA amongst cancer survivors. 29

Table S19. Predicted HR of All-cause mortality amongst Cancer survivors from Isotemporal substitution around sample average of MPA whilst holding other components constant at sample average. 30

Table S20. Predicted HR of Subsequent Cancer mortality amongst Cancer survivors from Isotemporal substitution around sample average of MPA whilst holding other components constant at sample average. 31

Table S21. Association of Coordinates of daily-movement behaviours (intensities, including MVPA) and sleep with risk of CVD-mortality among previous cancer survivors (Cox Proportional Hazards Model) excluding all self-reported history of cancer (n=2185), any cancer events within 2y prior to accelerometry measurement (n=481), and events within 2y of follow-up (n=27). 32

Table S22. Association of Coordinates of daily-movement behaviours (intensities, including MVPA) and sleep with risk of All-cause mortality among previous cancer survivors (Cox Proportional Hazards Model) excluding all self-reported history of cancer (n=2185), all cancer events within 2y prior to accelerometry measurement (n=481), and events within 2y follow-up (n=27). 33

Table S23. Association of Coordinates of daily-movement behaviours (intensities, including MVPA) and sleep with risk of cancer-mortality among previous cancer survivors (Cox Proportional Hazards Model) excluding all self-reported history of cancer (n=2185), all cancer events within 2y prior to accelerometry measurement (n=481), and events within 2y follow-up (n=27). 34

Tab**le S24**. Association of Coordinates of daily-movement behaviours (intensities, including MVPA) and sleep with risk of CVD-mortality among previous cancer survivors (Cox Proportional Hazards Model) excluding all self-reported history of cancer (n=2185) and all non-melanoma skin cancers (n=674) 35

Tab**le S25**. Association of Coordinates of daily-movement behaviours (intensities, including MVPA) and sleep with risk of Cancer-mortality among previous cancer survivors (Cox Proportional Hazards Model) excluding all self-reported history of cancer (n=2185) and all non-melanoma skin cancers (n=674) 36

Tab**le S26**. Association of Coordinates of daily-movement behaviours (intensities, including MVPA) and sleep with risk of All-cause mortality among previous cancer survivors (Cox Proportional Hazards Model) excluding all self-reported history of cancer (n=2185) and all non-melanoma skin cancers (n=674) 37

Table S27. E-Values (estimate of minimal association of a residual confounder with the exposure and outcome (All-cause mortality), required to nullify the associations observed in the fully-adjusted model). 38

Table S28. E-Values (estimate of minimal association of a residual confounder with the exposure and outcome (Cancer-mortality), required to nullify the associations observed in the fully-adjusted model). 38

Table S29. E-Values (estimate of minimal association of a residual confounder with the exposure and outcome (CVD-mortality), required to nullify the associations observed in the fully-adjusted model). 39

Supplementary references 40

Table S1a Cancer history ICD-10 codes.

| Cancer site | ICD-10 codes |
|---|---|
| Breast | C50 |
| Non-melanoma skin | C44 |
| Colorectal | C18–C21 |
| Blood | C81–C86, C88, C90–C96 |
| Prostate | C61 |
| Bladder | C67 |
| Melanoma | C43 |
| Other neoplasms | C46–C49, C51–C52, C57–C58, C60, C63, C76, C80, C97 |
| Uterus | C54–C55 |
| Lung | C33–C34 |
| Oesophagus | C15 |
| Kidney | C64 |
| Other | Other / unmatched ICD-10 codes |
| Ovary | C56 |
| Lip / oral cavity / pharynx | C01–C04 |
| Stomach | C16 |
| Endocrine gland | C73–C75 |
| Central nervous system | C69–C72 |
| Pancreas | C25 |
| Testis | C62 |
| Sinuses / larynx / trachea | C31–C33 |
| Bone & articular cartilage | C40–C41 |
| Cervix | C53 |
| Liver & intrahepatic bile ducts | C22 |
| Ill-defined digestive organs | C26 |
| Renal pelvis & ureter | C65–C66 |
| Small intestine | C17 |
| Other urinary organs | C68 |
| Gallbladder & biliary duct | C23–C24 |
| Mesothelioma | C45 |
| Ill-defined respiratory system | C30, C37, C39 |

Table S1b. Cancer sites of cancer survivor sample (excl. self-reported cases (n=2185)).

| **Primary cancer site** | **Cases** | **Percentage** |
|---|---|---|
| Breast cancer | 1298 | 32.7 |
| Non-melanoma skin cancer | 674 | 17 |
| Prostate cancer | 410 | 10.3 |
| Colorectal cancer | 369 | 9.3 |
| Bladder cancer | 157 | 4 |
| Melanoma | 153 | 3.9 |
| Endometrial cancer | 150 | 3.8 |
| Lymphoma | 140 | 3.5 |
| Leukaemia | 67 | 1.7 |
| Head and neck cancer | 66 | 1.7 |
| Secondary, ill-defined or unknown primary cancer | 57 | 1.4 |
| Kidney cancer | 54 | 1.4 |
| Ovarian cancer | 43 | 1.1 |
| Multiple myeloma | 42 | 1.1 |
| Lung cancer | 38 | 1 |
| Oesophageal cancer | 36 | 0.9 |
| Testicular cancer | 31 | 0.8 |
| Thyroid cancer | 31 | 0.8 |
| Soft tissue and retroperitoneal cancer | 24 | 0.6 |
| Cervical cancer | 21 | 0.5 |
| Laryngeal cancer | 17 | 0.4 |
| Stomach cancer | 15 | 0.4 |
| Eye cancer | 10 | 0.3 |
| Bone cancer | 9 | 0.2 |
| Anal cancer | 8 | 0.2 |
| Small intestinal cancer | 6 | 0.2 |
| Uterine cancer, unspecified | 6 | 0.2 |
| Other digestive organ cancer | 5 | 0.1 |
| Other cancers* | 36 | 0.9 |

*Bin widths <5 cases repressed according to UKBB disclosure policy. Other cancer sites include: Penile cancer, haematological malignancies, pancreatic cancer, vulval cancer, brain/CNS cancer, liver cancer, renal/ureter cancer, nasal/sinus cancer, urinary tract cancer, adrenal cancer, biliary tract cancer, other cancers, female genital cancers, male genital cancers, and thoracic cancers.

### Supplementary text S1. Sleep and Physical activity intensity classification algorithms.

#### Sleep classifier

Sleep duration was assessed with an algorithm, using the absence of change in device tilt angle, developed and validated in 3,752 British participants with sleep diary data and 28 patients with polysomnography data. The sleep period time window derived from the algorithm was 11m and 3m longer compared with a sleep diary in men and women, respectively. The mean c-statistic to detect the sleep period time window compared to polysomnography was 0.86 and 0.83 in clinic-based and healthy sleepers, respectively[1].

#### Physical activity intensity classifier

Physical activity intensity was classified with a previously adopted two-level Random Forest algorithm[2, 3]. The first level classifies activity type, in contiguous 10s bins, and the second level further sorts the activities into intensity bands. This two-level classifier minimises misclassification between the lower intensity bands as activities are first classified by type, clearly delineating Moving activities, standing utilitarian movements (semi-stationary) and sedentary behaviour (SB) before non-sedentary activities are further classified into intensity bands based on degree of acceleration against gravity. Walking activities only are then grouped by their acceleration, as light PA (LPA; <100mg), moderate PA (MPA; ≥100mg) and vigorous (≥400mg). The intensity composition therefore encompassed VPA, MPA, LPA, sedentary time (ST) and sleep.

### Supplementary text S2. Compositional 'pivot' approach

The pivot approach is an established approach in compositional data analysis[4, 5] enables the joint modelling of highly collinear movement behaviours by applying the isometric log-ratio (ILR) transformation to compositions of $n$ components. This yields $n$–1 ILR coordinates (ratios), which collectively represent the full 24-hour movement composition and can be included simultaneously in standard regression models, thereby avoiding multicollinearity[6, 7].

In each model, the first ILR coordinate represents the association of a single movement behaviour relative to the remaining behaviours (e.g. MVPA relative to LPA, sedentary behaviour, and sleep) with the outcome. Only this first ILR coefficient is interpreted, corresponding to the expected change in the outcome per one-unit increase in the coordinate. The composition is then re-pivoted and the model re-estimated so that each movement behaviour is, in turn, expressed relative to all others.

Given the difficulty in interpreting ILR-transformed units, isotemporal substitution was performed to understand the estimated change in event hazard from a minute-by-minute reallocation of time from one (significant) behaviour into each other behaviour, around the sample average.

As log-ratio transformation requires all components to be non-zero (<0.2% of the sample accrued an average of 0 minutes of VPA across the wear week). Zero values also censor data below the minimal detection threshold of the device. Thus, to impute zeroes with the minimum detection limit of the device, a log-ratio expectation-likelihood (LREM) multiplicative zero replacement algorithm, from zCompositions, was used.

All analyses were performed in R version 4.4.3.

Table S2 Included Covariate definitions.

| Variable | Coding | UKB field |
| --- | --- | --- |
| Age | Continuous (years) | 34, 52, accelerometer date-timestamp |
| Sex | Female/Male | 31 |
| Smoker status | Never, past, current smoker. | 20116 |
| Alcohol Use | Never drinker, Ex-drinker Within guidelines and 4 units/wk, Above guidelines and | 20117, 1558 |
| Diet | Fruits and vegetables servings/day | 1309, 1319, 1289, 1299 |
| Prevalent CVD | Yes/No Identified by self-report and hospitalisation (ICD-10: I0, I11, I13, I20-I51, I60-I69). | 20002, 41259 |
| Education | College/University; A/AS level; O levels; CSE; NVQ/HND/HNC; other. A/AS = Advanced Placement; O level = High school certificate; CSE = Certificate of secondary education; NVQ/HND/HNC = Vocational qualification / Associates degree. A level, typically at age 18 years; O level, typically at age 16 years; CSE, typically at age 16 years; | 6138 |
| cholesterol medication | Yes/No | 6177, 6153 |
| blood pressure medication | Yes/No | 6177, 6153 |
| Use of diabetes medication | Yes/No | 6177, 6153 |
| Parental history of cancer | Yes/No Self-reported mother or father diagnosed with cancer | 20107, 20110 |

Figure S1 Sample derivation for UK Biobank participant inclusion resulting in n=6,158 included participants.

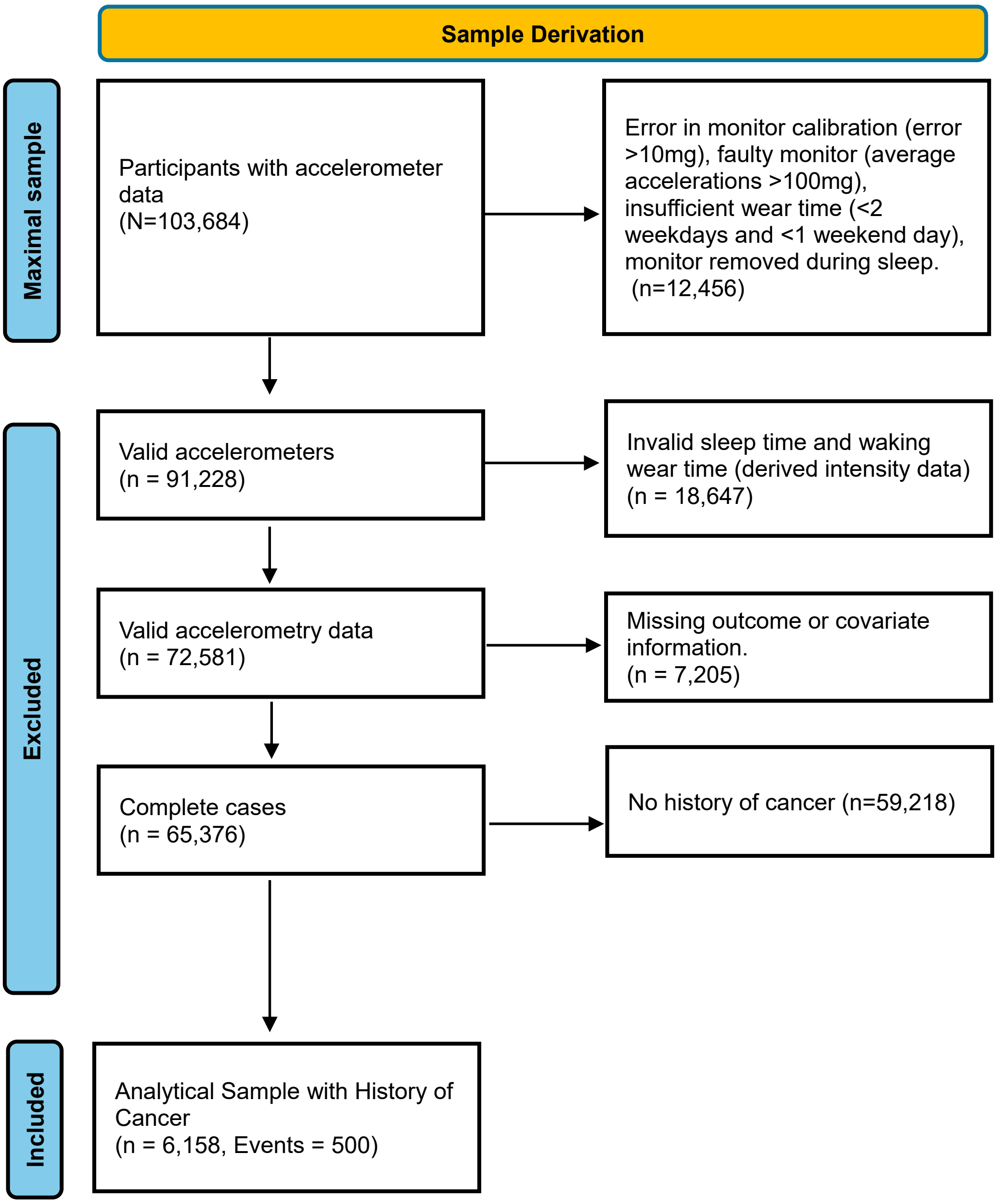

Table S3. Movement behaviour, and difference from the sample compositional average, by outcome.

| Outcome | Behaviour | Average mins | % Difference | Lower CI% | Upper CI% | Lower CI (mins) | Upper CI (mins) |
|---|---|---|---|---|---|---|---|
| **Sample centre (Ref)** | **MVPA** | **27** | - | - | - | - | - |
| | **LPA** | **297** | - | - | - | - | - |
| | **SB** | **653** | - | - | - | - | - |
| | **Sleep** | **463** | - | - | - | - | - |
| **All-cause mortality** | **MVPA** | **18** | -43 | -51 | -35 | -14 | -10 |
| | **LPA** | **282** | -5 | -7 | -4 | -21 | -10 |
| | **SB** | **688** | 5 | 4 | 6 | 27 | 42 |
| | **Sleep** | **452** | -2 | -4 | -1 | -18 | -3 |
| **CVD-mortality** | **MVPA** | **16** | -53 | -69 | -36 | -19 | -10 |
| | **LPA** | **277** | -7 | -11 | -3 | -32 | -9 |
| | **SB** | **709** | 8 | 6 | 11 | 38 | 70 |
| | **Sleep** | **438** | -6 | -9 | -2 | -42 | -8 |
| **Cancer-mortality** | **MVPA** | **19** | -38 | -47 | -28 | -13 | -8 |
| | **LPA** | **284** | -4 | -7 | -2 | -19 | -7 |
| | **SB** | **683** | 5 | 3 | 6 | 20 | 38 |
| | **Sleep** | **454** | -2 | -4 | 0 | -17 | 0 |

Percentage (%) difference from compositional centre & 95% Confidence intervals (CI) estimated by 1000 bootstrapped samples.

Table S4. Movement behaviour (separated by intensity), and difference from the sample compositional average, by outcome.

| Outcome | Behaviour | Average mins | % Difference | Diff. Lower CI% | Diff. Upper CI% | Diff. Lower CI (mins) | Diff. Upper CI (mins) |
|---|---|---|---|---|---|---|---|
| **Sample centre (Ref)** | **VPA** | **2** | | | | | |
| | **MPA** | **25** | - | - | - | - | - |
| | **LPA** | **297** | - | - | - | - | - |
| | **SB** | **652** | - | - | - | - | - |
| | **Sleep** | **463** | - | - | - | - | - |
| **All-cause mortality** | **VPA** | **1** | -44 | -38 | -50 | -1 | -1 |
| | **MPA** | **16** | -34 | -29 | -39 | -8 | -7 |
| | **LPA** | **282** | -5 | -4 | -7 | -15 | -10 |
| | **SB** | **689** | 5 | 7 | 4 | 35 | 43 |
| | **Sleep** | **452** | -2 | -1 | -4 | -11 | -4 |
| **CVD-mortality** | **VPA** | **1** | -53 | -41 | -63 | -1 | -1 |
| | **MPA** | **15** | -40 | -29 | -49 | -10 | -7 |
| | **LPA** | **277** | -7 | -3 | -10 | -20 | -9 |
| | **SB** | **709** | 9 | 11 | 6 | 56 | 74 |
| | **Sleep** | **439** | -5 | -2 | -9 | -25 | -9 |
| **Cancer-mortality** | **VPA** | **2** | -37 | -28 | -45 | -1 | -1 |
| | **MPA** | **17** | -30 | -23 | -37 | -7 | -6 |
| | **LPA** | **284** | -4 | -2 | -6 | -13 | -7 |
| | **SB** | **683** | 5 | 6 | 3 | 30 | 39 |
| | **Sleep** | **454** | -2 | 0 | -4 | -9 | 0 |

Percentage (%) difference from compositional centre & 95% Confidence intervals (CI) estimated by 1000 bootstrapped samples. Difference: Diff

Figure S2. Relative differences in 24-hour time use (separated by intensity) by mortality status (compared to cancer survivors with no mortality event).

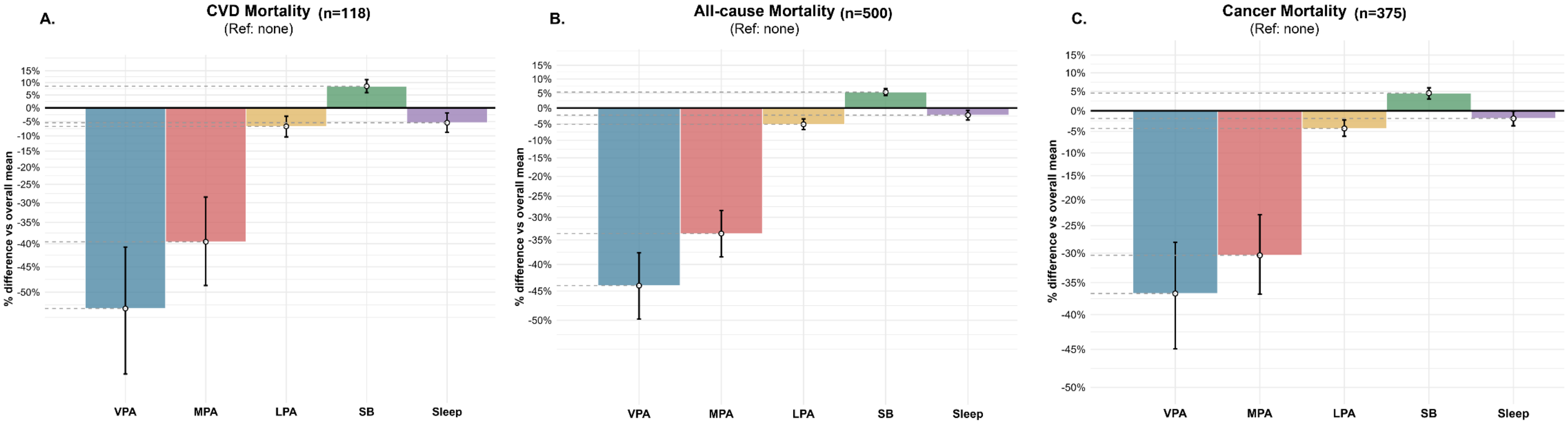

Table S5. Association of Coordinates of daily-movement behaviours (intensities, including MVPA) and sleep with risk of CVD-mortality among previous cancer survivors (Fine-Gray Cox Proportional Hazards Model)

| | Age and Sex-adjusted | | | | Fully adjusted | | | |
|---|---|---|---|---|---|---|---|---|
| | **HR** | **95% CI** | | **p-value** | **HR** | **95% CI** | | **p-value** |
| **Moderate-to-Vigorous PA** (vs all other behaviours) | **0.79** | **0.71** | **0.88** | **<0.001** | **0.82** | **0.73** | **0.92** | **<0.001** |
| Light PA (vs all other behaviours) | 0.95 | 0.63 | 1.44 | 0.824 | 0.99 | 0.66 | 1.54 | 0.979 |
| **Sleep** (vs all other behaviours) | **0.63** | **0.45** | **0.88** | **0.008** | **0.65** | **0.46** | **0.92** | **0.016** |
| **Sedentary time** (vs all other behaviours) | **2.11** | **1.40** | **3.18** | **0.004** | **1.87** | **1.23** | **2.84** | **0.004** |

**Coefficients are ILR transformed units and not directly interpretable.**

Table S6. Association of Coordinates of daily-movement behaviours (intensities, including MVPA) and sleep with risk of All-cause mortality among previous cancer survivors (Cox Proportional Hazards Model)

| | **Age and Sex-adjusted** | | | | **Fully adjusted** | | | |
|---|---|---|---|---|---|---|---|---|
| | **HR** | **95% CI** | | **p-value** | **HR** | **95% CI** | | **p-value** |
| **Moderate-to-Vigorous PA** (vs all other behaviours) | **0.79** | **0.74** | **0.83** | **<0.001** | **0.79** | **0.75** | **0.84** | **<0.001** |
| Light PA (vs all other behaviours) | 1.02 | 0.84 | 1.26 | 0.816 | 1.05 | 0.85 | 1.28 | 0.649 |
| Sleep (vs all other behaviours) | 0.85 | 0.70 | 1.03 | 0.091 | 0.86 | 0.72 | 1.05 | 0.145 |
| **Sedentary time** (vs all other behaviours) | **1.48** | **1.20** | **1.80** | **<0.001** | **1.39** | **1.13** | **1.70** | **0.002** |

**Coefficients are ILR transformed units and not directly interpretable.**

Table S7. Association of Coordinates of daily-movement behaviours (intensities, including MVPA) and sleep with risk of cancer-mortality among previous cancer survivors (Fine-Gray Cox Proportional Hazards Model)

| | **Age and Sex-adjusted** | | | | **Fully adjusted** | | | |
|---|---|---|---|---|---|---|---|---|
| | **HR** | **95% CI** | | **p-value** | **HR** | **95% CI** | | **p-value** |
| **Moderate-to-Vigorous PA** (vs all other behaviours) | **0.80** | **0.75** | **0.85** | **<0.001** | **0.81** | **0.76** | **0.87** | **<0.001** |
| Light PA (vs all other behaviours) | 1.04 | 0.83 | 1.32 | 0.731 | 1.06 | 0.84 | 1.34 | 0.616 |
| Sleep (vs all other behaviours) | 0.88 | 0.70 | 1.10 | 0.260 | 0.89 | 0.72 | 1.12 | 0.313 |
| **Sedentary time** (vs all other behaviours) | **1.36** | **1.07** | **1.73** | **0.011** | **1.30** | **1.02** | **1.65** | **0.035** |

**Coefficients are ILR transformed units and not directly interpretable.**

Table S8. Predicted HR of CVD-mortality amongst Cancer survivors from Isotemporal substitution around sample average of MVPA whilst holding other components constant at sample average.

| | | MVPA → SB | | | | MVPA → SLEEP | | | | MVPA → LPA | | | |
|---|---|---|---|---|---|---|---|---|---|---|---|---|---|
| | **ΔRisk** | **MVPA (daily mins)** | **HR** | **95% CI** | | **MVPA (daily mins)** | **HR** | **95% CI** | | **MVPA (daily mins)** | **HR** | **95% CI** | |
| **Decreasing MVPA** | **25%** | 17 | **1.24** | 1.11 | 1.38 | 15 | **1.24** | 1.08 | 1.44 | 16 | **1.24** | 1.07 | 1.44 |
| | **20%** | 18 | **1.21** | 1.10 | 1.33 | 17 | **1.19** | 1.06 | 1.33 | 18 | **1.19** | 1.06 | 1.33 |
| | **15%** | 20 | **1.16** | 1.08 | 1.24 | 19 | **1.14** | 1.05 | 1.25 | 20 | **1.14** | 1.04 | 1.25 |
| | **10%** | 22 | **1.11** | 1.05 | 1.17 | 21 | **1.10** | 1.03 | 1.18 | 22 | **1.10** | 1.03 | 1.17 |
| | **5%** | 25 | **1.05** | 1.03 | 1.08 | 24 | **1.05** | 1.02 | 1.09 | 25 | **1.04** | 1.01 | 1.08 |
| **Increasing MVPA** | **-5%** | 31 | **0.96** | 0.94 | 0.98 | 33 | **0.95** | 0.91 | 0.98 | 32 | **0.95** | 0.91 | 0.99 |
| | **-10%** | 35 | **0.91** | 0.86 | 0.95 | 39 | **0.90** | 0.83 | 0.97 | 37 | **0.90** | 0.83 | 0.97 |
| | **-15%** | 40 | **0.85** | 0.79 | 0.92 | 47 | **0.85** | 0.75 | 0.96 | 43 | **0.85** | 0.75 | 0.96 |
| | **-20%** | 46 | **0.80** | 0.72 | 0.89 | 58 | **0.80** | 0.67 | 0.95 | 50 | **0.80** | 0.67 | 0.95 |
| | **-25%** | 52 | **0.75** | 0.66 | 0.86 | 74 | **0.75** | 0.59 | 0.95 | 59 | **0.75** | 0.60 | 0.94 |

Figure S3. Difference in hazard of subsequent All-cause mortality from theoretical reallocation of time into/out of MVPA amongst cancer survivors.

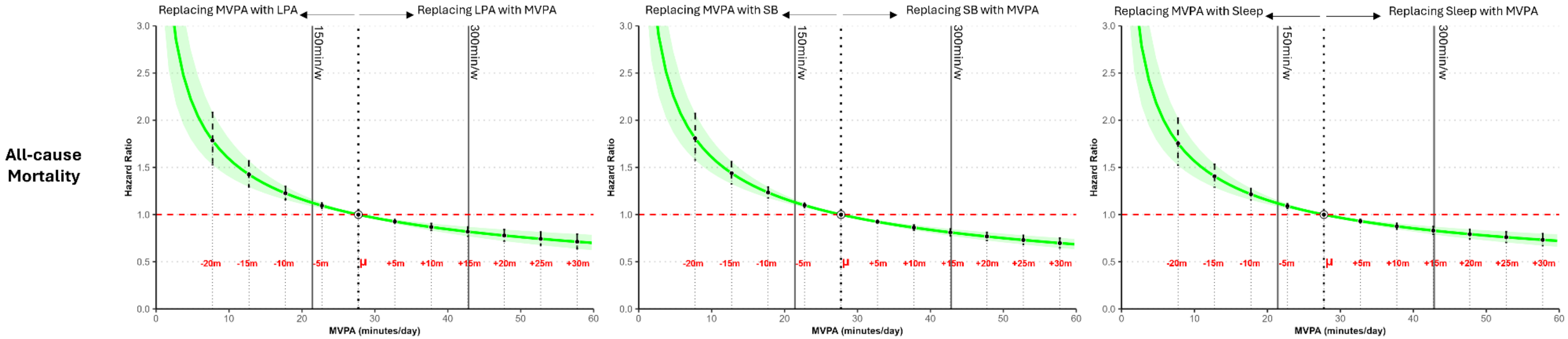


Table S9. Predicted HR of All-cause mortality amongst Cancer survivors from Isotemporal substitution around sample average of MVPA whilst holding other components constant at sample average.

| | ΔRisk | MVPA → SB<br>MVPA (daily mins) | HR | 95% CI | | MVPA → SLEEP<br>MVPA (daily mins) | HR | 95% CI | | MVPA → LPA<br>MVPA (daily mins) | HR | 95% CI | |
|---|---|---|---|---|---|---|---|---|---|---|---|---|---|
| **Decreasing MVPA** | **25%** | 18 | **1.23** | 1.18 | 1.29 | 17 | **1.25** | 1.18 | 1.32 | 17 | **1.26** | 1.18 | 1.34 |
| | **20%** | 19 | **1.20** | 1.15 | 1.25 | 19 | **1.19** | 1.14 | 1.24 | 19 | **1.20** | 1.14 | 1.26 |
| | **15%** | 21 | **1.15** | 1.11 | 1.18 | 20 | **1.16** | 1.12 | 1.21 | 21 | **1.14** | 1.10 | 1.19 |
| | **10%** | 23 | **1.10** | 1.08 | 1.12 | 23 | **1.09** | 1.07 | 1.11 | 23 | **1.10** | 1.07 | 1.12 |
| | **5%** | 25 | **1.06** | 1.04 | 1.07 | 25 | **1.05** | 1.04 | 1.06 | 25 | **1.05** | 1.04 | 1.07 |
| **Increasing MVPA** | **-5%** | 31 | **0.95** | 0.94 | 0.96 | 32 | **0.94** | 0.93 | 0.96 | 31 | **0.95** | 0.94 | 0.97 |
| | **-10%** | 35 | **0.90** | 0.88 | 0.92 | 36 | **0.90** | 0.87 | 0.92 | 35 | **0.90** | 0.87 | 0.93 |
| | **-15%** | 39 | **0.85** | 0.82 | 0.88 | 41 | **0.85** | 0.81 | 0.89 | 40 | **0.85** | 0.81 | 0.89 |
| | **-20%** | 44 | **0.80** | 0.77 | 0.84 | 47 | **0.80** | 0.75 | 0.85 | 45 | **0.80** | 0.75 | 0.86 |
| | **-25%** | 50 | **0.75** | 0.71 | 0.80 | 55 | **0.75** | 0.69 | 0.81 | 52 | **0.75** | 0.68 | 0.82 |

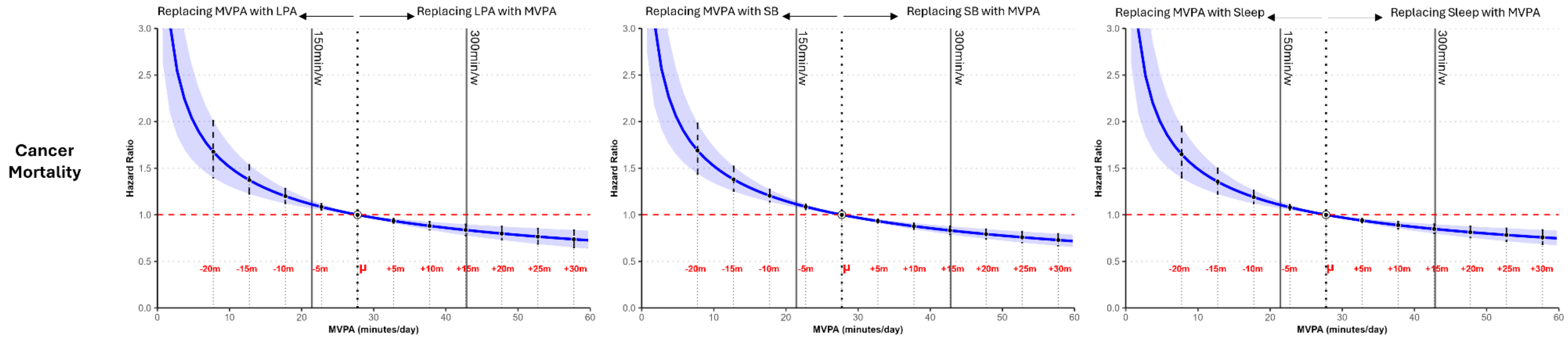


Figure S4. Difference in hazard of subsequent cancer-mortality from theoretical reallocation of time into/out of MVPA amongst cancer survivors.

Table S10. Predicted HR of Cancer-specific mortality amongst former Cancer survivors from Isotemporal substitution around sample average of MVPA whilst holding other components constant at sample average.

| | | **MVPA** | | **→ SB** | | **MVPA** | | **→ SLEEP** | | **MVPA** | | **→ LPA** | |
|---|---|---|---|---|---|---|---|---|---|---|---|---|---|
| | **ΔRisk** | **(daily mins)** | **HR** | **95% CI** | | **(daily mins)** | **HR** | **95% CI** | | **(daily mins)** | **HR** | **95% CI** | |
| **Decreasing MVPA** | **25%** | 16 | **1.27** | 1.18 | 1.36 | 16 | **1.25** | 1.16 | 1.34 | 16 | **1.26** | 1.16 | 1.37 |
| | **20%** | 18 | **1.20** | 1.14 | 1.27 | 18 | **1.19** | 1.12 | 1.26 | 18 | **1.20** | 1.12 | 1.28 |
| | **15%** | 20 | **1.15** | 1.11 | 1.20 | 20 | **1.14** | 1.09 | 1.19 | 20 | **1.15** | 1.09 | 1.21 |
| | **10%** | 22 | **1.11** | 1.07 | 1.14 | 22 | **1.10** | 1.06 | 1.14 | 22 | **1.10** | 1.06 | 1.15 |
| | **5%** | 25 | **1.05** | 1.03 | 1.06 | 25 | **1.05** | 1.03 | 1.06 | 25 | **1.05** | 1.03 | 1.07 |
| **Increasin g MVPA** | **-5%** | 32 | **0.94** | 0.93 | 0.96 | 32 | **0.95** | 0.93 | 0.97 | 32 | **0.95** | 0.93 | 0.97 |
| | **-10%** | 36 | **0.90** | 0.87 | 0.93 | 37 | **0.90** | 0.86 | 0.93 | 36 | **0.90** | 0.87 | 0.94 |
| | **-15%** | 41 | **0.85** | 0.81 | 0.89 | 43 | **0.85** | 0.80 | 0.90 | 41 | **0.85** | 0.80 | 0.91 |
| | **-20%** | 47 | **0.80** | 0.75 | 0.85 | 50 | **0.80** | 0.74 | 0.87 | 48 | **0.80** | 0.73 | 0.87 |

| | | | | | | | | | | | | | |
|---|---|---|---|---|---|---|---|---|---|---|---|---|---|
| | **-25%** | 54 | **0.75** | 0.69 | 0.82 | 60 | **0.75** | 0.67 | 0.83 | 56 | **0.75** | 0.66 | 0.84 |

Table S11. Predicted HR of CVD-mortality amongst Cancer survivors from Isotemporal substitution around sample average of Sleep whilst holding other components constant at sample average.

| | ΔRisk | Sleep (daily mins) | HR | → SB 95% CI | |
|---|---|---|---|---|---|
| Decreasing Sleep | **25%** | 403 | **1.25** | 1.08 | 1.45 |
| | **20%** | 414 | **1.20** | 1.06 | 1.35 |
| | **15%** | 425 | **1.15** | 1.05 | 1.26 |
| | **10%** | 437 | **1.10** | 1.03 | 1.17 |
| | **5%** | 450 | **1.05** | 1.02 | 1.08 |
| Increasing Sleep | **-5%** | 477 | **0.95** | 0.92 | 0.98 |
| | **-10%** | 492 | **0.90** | 0.84 | 0.96 |
| | **-15%** | 508 | **0.85** | 0.76 | 0.94 |
| | **-20%** | 524 | **0.80** | 0.69 | 0.93 |
| | **-25%** | 542 | **0.75** | 0.62 | 0.90 |

Table S12. Predicted HR of All-cause mortality amongst Cancer survivors from Isotemporal substitution around sample average of Sleep whilst holding other components constant at sample average.

| | ΔRisk | Sleep (daily mins) | HR | → SB 95% CI | |
|---|---|---|---|---|---|
| **Decreasing Sleep** | **25%** | 321 | **1.25** | 1.03 | 1.52 |
| | **20%** | 347 | **1.20** | 1.03 | 1.40 |
| | **15%** | 373 | **1.15** | 1.02 | 1.29 |
| | **10%** | 402 | **1.10** | 1.02 | 1.19 |
| | **5%** | 432 | **1.05** | 1.01 | 1.09 |
| **Increasing Sleep** | **-5%** | 496 | **0.95** | 0.91 | 0.99 |
| | **-10%** | 530 | **0.90** | 0.83 | 0.98 |
| | **-15%** | 566 | **0.85** | 0.75 | 0.97 |
| | **-20%** | 602 | **0.80** | 0.67 | 0.95 |
| | **-25%** | 640 | **0.75** | 0.60 | 0.93 |

Table S13. Association of Coordinates of daily-movement behaviours (intensities) and sleep with risk of CVD-mortality (Fine-Gray Cox Proportional Hazards Model)

| | Age and Sex-adjusted | | | | Fully-adjusted | | | |
|---|---|---|---|---|---|---|---|---|
| | HR | 95% CI | | p-value | HR | 95% CI | | p-value |
| **Vigorous physical activity** (vs all other postures) | **0.72** | **0.59** | **0.89** | **<0.001** | **0.76** | **0.62** | **0.93** | **<0.001** |
| **Moderate physical activity** (vs all other postures) | 0.76 | 0.58 | 1.00 | 0.054 | 0.80 | 0.60 | 1.06 | 0.117 |
| **Light physical activity** (vs all other postures) | 0.85 | 0.35 | 2.07 | 0.726 | 0.96 | 0.39 | 2.37 | 0.932 |
| **Sedentary behaviour** (vs all other postures) | **4.86** | **1.99** | **11.81** | **<0.001** | **3.76** | **1.52** | **9.29** | **0.004** |
| **Sleep duration** (vs all other postures) | **0.40** | **0.19** | **0.83** | **0.014** | **0.42** | **0.20** | **0.90** | **0.025** |

**Coefficients are ILR transformed units and not directly interpretable.**

Table S14. Association of Coordinates of daily-movement behaviours (intensities) and sleep with risk of All-cause mortality (Fine-Gray Cox Proportional Hazards Model)

| | **Age and Sex-adjusted** | | | | **Fully-adjusted** | | | |
|---|---|---|---|---|---|---|---|---|
| | **HR** | **95% CI** | | **p-value** | **HR** | **95% CI** | | **p-value** |
| **Vigorous physical activity** (vs all other postures) | **0.77** | **0.70** | **0.85** | **<0.001** | **0.79** | **0.71** | **0.87** | **<0.001** |
| **Moderate physical activity** (vs all other postures) | **0.77** | **0.67** | **0.90** | **<0.001** | **0.77** | **0.67** | **0.90** | **<0.001** |
| **Light physical activity** (vs all other postures) | 1.01 | 0.65 | 1.56 | 0.963 | 1.06 | 0.68 | 1.64 | 0.801 |
| **Sedentary behaviour** (vs all other postures) | **2.28** | **1.46** | **3.55** | **<0.001** | **2.01** | **1.28** | **3.15** | **0.002** |
| **Sleep duration** (vs all other postures) | 0.74 | 0.50 | 1.11 | 0.145 | 0.77 | 0.51 | 1.16 | 0.208 |

**Coefficients are ILR transformed units and not directly interpretable.**

Table S15. Association of Coordinates of daily-movement behaviours (intensities) and sleep with risk of cancer-mortality (Fine-Gray Cox Proportional Hazards Model)

| | Age and Sex-adjusted | | | | Fully-adjusted | | | |
|---|---|---|---|---|---|---|---|---|
| | HR | 95% CI | | p-value | HR | 95% CI | | p-value |
| **Vigorous physical activity**<br>(vs all other postures) | **0.85** | **0.76** | **0.96** | **0.006** | **0.87** | **0.77** | **0.98** | **0.021** |
| **Moderate physical activity**<br>(vs all other postures) | **0.73** | **0.62** | **0.87** | **<0.001** | **0.74** | **0.63** | **0.88** | **<0.001** |
| **Light physical activity**<br>(vs all other postures) | 1.06 | 0.64 | 1.75 | 0.8298 | 1.10 | 0.66 | 1.83 | 0.710 |
| **Sedentary behaviour**<br>(vs all other postures) | **1.95** | **1.16** | **3.27** | **0.012** | **1.76** | **1.05** | **2.97** | **0.034** |
| **Sleep duration**<br>(vs all other postures) | 0.78 | 0.49 | 1.26 | 0.3081 | 0.80 | 0.50 | 1.28 | 0.352 |

**Coefficients are ILR transformed units and not directly interpretable.**

Figure S5. Difference in hazard of all mortality outcomes from theoretical reallocation of time into/out of VPA amongst cancer survivors.

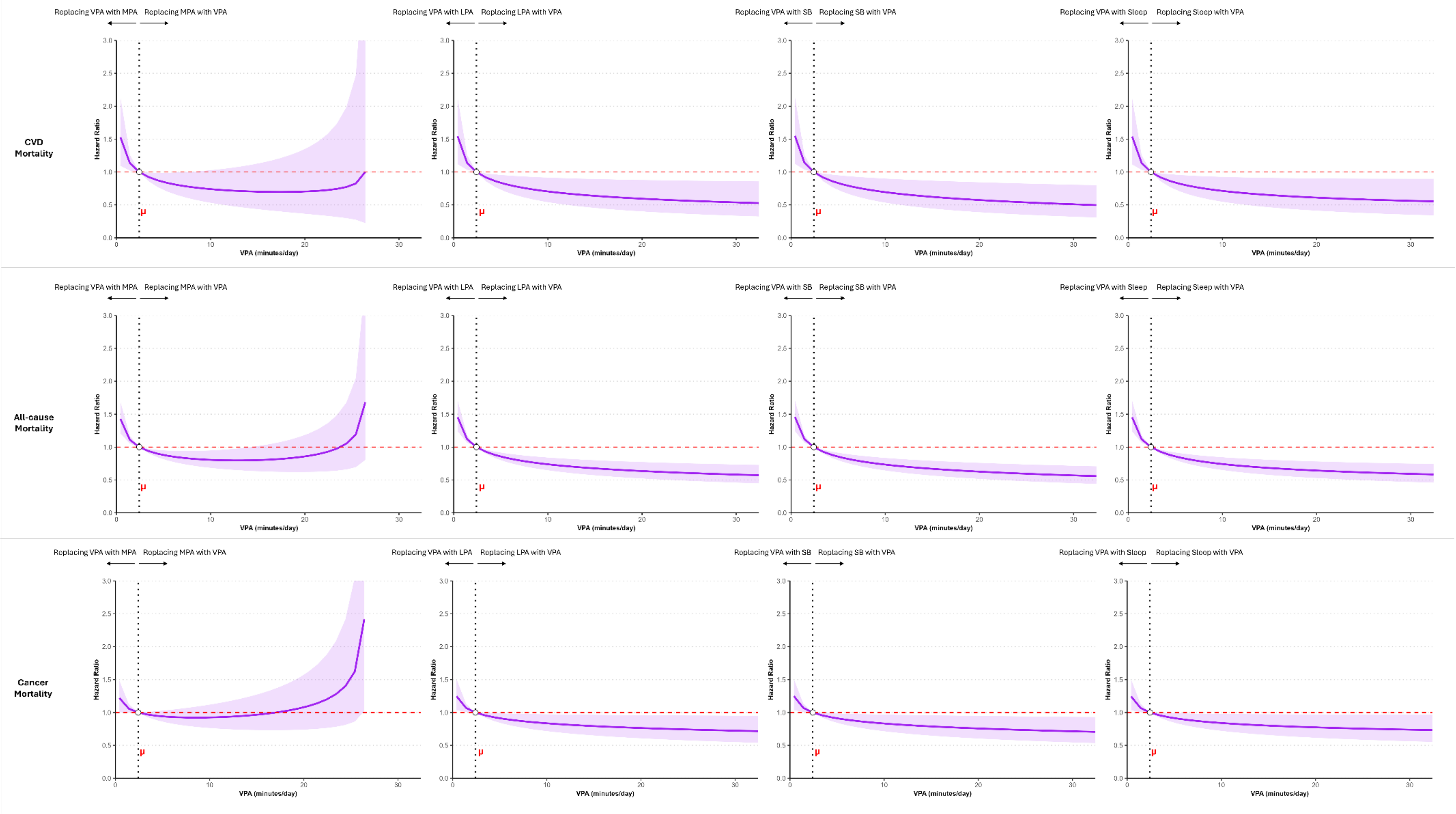

Table S16. Predicted HR of CVD-mortality amongst Cancer survivors from Isotemporal substitution around sample average of VPA whilst holding other components constant at sample average.

| | ΔRisk | VPA (daily mins) | → SB HR | 95% CI | | VPA (daily mins) | → MPA HR | 95% CI | | VPA (daily mins) | → SLEEP HR | 95% CI | | VPA (daily mins) | → LPA HR | 95% CI | |
|---|---|---|---|---|---|---|---|---|---|---|---|---|---|---|---|---|---|
| **Decreasing VPA** | **25%** | - | **-** | - | - | - | - | **-** | - | - | - | - | **-** | - | - | - | - |
| | **20%** | - | **-** | - | - | - | - | **-** | - | - | - | - | **-** | - | - | - | - |
| | **15%** | 1 | **1.14** | 1.04 | 1.26 | 1 | **1.14** | 1.02 | 1.26 | 1 | **1.14** | 1.03 | 1.26 | 21 | **1.14** | 1.03 | 1.26 |
| | **10%** | - | **-** | - | - | - | - | **-** | - | - | - | - | **-** | - | - | - | - |
| | **5%** | - | **-** | - | - | - | - | **-** | - | - | - | - | **-** | - | - | - | - |
| **Increasing VPA** | **-5%** | - | **-** | - | - | 3 | **-** | - | - | 3 | **-** | - | - | - | **-** | - | - |
| | **-10%** | 3 | **0.92** | 0.86 | 0.98 | 3 | **0.92** | 0.86 | 0.99 | 3 | **0.92** | 0.86 | 0.98 | 3 | **0.92** | 0.86 | 0.98 |
| | **-15%** | 4 | **0.86** | 0.77 | 0.96 | 5 | **0.83** | 0.70 | 0.99 | 4 | **0.86** | 0.77 | 0.97 | 4 | **0.86** | 0.77 | 0.96 |
| | **-20%** | 5 | **0.81** | 0.70 | 0.94 | 6 | **0.80** | 0.65 | 0.99 | 6 | **0.79** | 0.66 | 0.95 | 6 | **0.79** | 0.66 | 0.94 |
| | **-25%** | 7 | **0.75** | 0.61 | 0.92 | 7 | **0.78** | 0.61 | 1.00 | 8 | **0.74** | 0.59 | 0.93 | 7 | **0.76** | 0.62 | 0.93 |

Table S17. Predicted HR of All-cause mortality amongst Cancer survivors from Isotemporal substitution around sample average of VPA whilst holding other components constant at sample average.

| | | VPA | → SB | | | VPA | → MPA | | | VPA | → SLEEP | | | VPA | → LPA | | |
|---|---|---|---|---|---|---|---|---|---|---|---|---|---|---|---|---|---|
| | **ΔRisk** | **(daily mins)** | **HR** | **95% CI** | | **(daily mins)** | **HR** | **95% CI** | | **(daily mins)** | **HR** | **95% CI** | | **(daily mins)** | **HR** | **95% CI** | |
| **Decreasing VPA** | **25%** | - | - | - | - | - | - | - | - | - | - | - | - | - | - | - | - |
| | **20%** | - | - | - | - | - | - | - | - | - | - | - | - | - | - | - | - |
| | **15%** | - | - | - | - | - | - | - | - | - | - | - | - | - | - | - | - |
| | **10%** | 1 | **1.12** | 1.07 | 1.18 | 1 | **1.11** | 1.05 | 1.17 | 1 | **1.12** | 1.07 | 1.18 | 1 | **1.12** | 1.07 | 1.18 |
| | **5%** | - | - | - | - | - | - | - | - | - | - | - | - | - | - | - | - |
| **Increasing VPA** | **-5%** | 3 | **0.93** | 0.90 | 0.96 | 3 | **0.94** | 0.91 | 0.97 | 3 | **0.93** | 0.90 | 0.96 | 3 | **0.93** | 0.90 | 0.96 |
| | **-10%** | 4 | **0.88** | 0.83 | 0.93 | 4 | **0.90** | 0.84 | 0.95 | 4 | **0.88** | 0.83 | 0.93 | 4 | **0.88** | 0.83 | 0.93 |
| | **-15%** | 5 | **0.84** | 0.78 | 0.90 | 6 | **0.85** | 0.76 | 0.94 | 5 | **0.84** | 0.78 | 0.91 | 5 | **0.84** | 0.78 | 0.91 |
| | **-20%** | 6 | **0.81** | 0.74 | 0.88 | 11 | **0.80** | 0.67 | 0.96 | 7 | **0.79** | 0.71 | 0.87 | 6 | **0.81** | 0.74 | 0.89 |
| | **-25%** | 9 | **0.74** | 0.66 | 0.84 | 12 | **0.80** | 0.65 | 0.97 | 9 | **0.75** | 0.66 | 0.85 | 9 | **0.75** | 0.66 | 0.85 |

Table S18. Predicted HR of subsequent cancer mortality amongst Cancer survivors from Isotemporal substitution around sample average of VPA whilst holding other components constant at sample average.

| | | VPA | → SB | | | VPA | → MPA | | | VPA | → SLEEP | | | VPA | → LPA | | |
|---|---|---|---|---|---|---|---|---|---|---|---|---|---|---|---|---|---|
| | **ΔRisk** | **(daily mins)** | **HR** | **95% CI** | | **(daily mins)** | **HR** | **95% CI** | | **(daily mins)** | **HR** | **95% CI** | | **(daily mins)** | **HR** | **95% CI** | |
| **Decreasing VPA** | **25%** | 0 | **1.25** | 1.03 | 1.50 | - | **-** | - | - | 0 | **1.24** | 1.03 | 1.50 | 0 | **1.25** | 1.03 | 1.50 |
| | **20%** | - | **-** | - | - | 0 | **1.22** | 1.00 | 1.48 | - | **-** | - | - | - | **-** | - | - |
| | **15%** | - | **-** | - | - | - | **-** | - | - | - | **-** | - | - | - | **-** | - | - |
| | **10%** | - | **-** | - | - | - | **-** | - | - | - | **-** | - | - | - | **-** | - | - |
| | **5%** | 1 | **1.07** | 1.01 | 1.13 | - | **-** | | | 1 | **1.07** | 1.01 | 1.13 | 1 | **1.07** | 1.01 | 1.13 |
| **Increasing VPA** | **-5%** | 3 | **0.96** | 0.92 | 0.99 | - | **-** | - | - | 3 | **0.96** | 0.92 | 0.99 | 3 | **0.96** | 0.92 | 0.99 |
| | **-10%** | 5 | **0.90** | 0.83 | 0.98 | - | **-** | - | - | 5 | **0.91** | 0.83 | 0.99 | 5 | **0.90** | 0.83 | 0.98 |
| | **-15%** | 8 | **0.85** | 0.75 | 0.97 | - | **-** | - | - | 9 | **0.85** | 0.73 | 0.98 | 8 | **0.85** | 0.75 | 0.98 |
| | **-20%** | 13 | **0.80** | 0.67 | 0.96 | - | **-** | - | - | 15 | **0.80** | 0.65 | 0.97 | 14 | **0.80** | 0.66 | 0.97 |
| | **-25%** | 21 | **0.75** | 0.60 | 0.95 | - | **-** | - | - | 26 | **0.75** | 0.58 | 0.97 | 23 | **0.75** | 0.59 | 0.96 |

Figure S6. Difference in hazard of all mortality outcomes from theoretical reallocation of time into/out of MPA amongst cancer survivors.

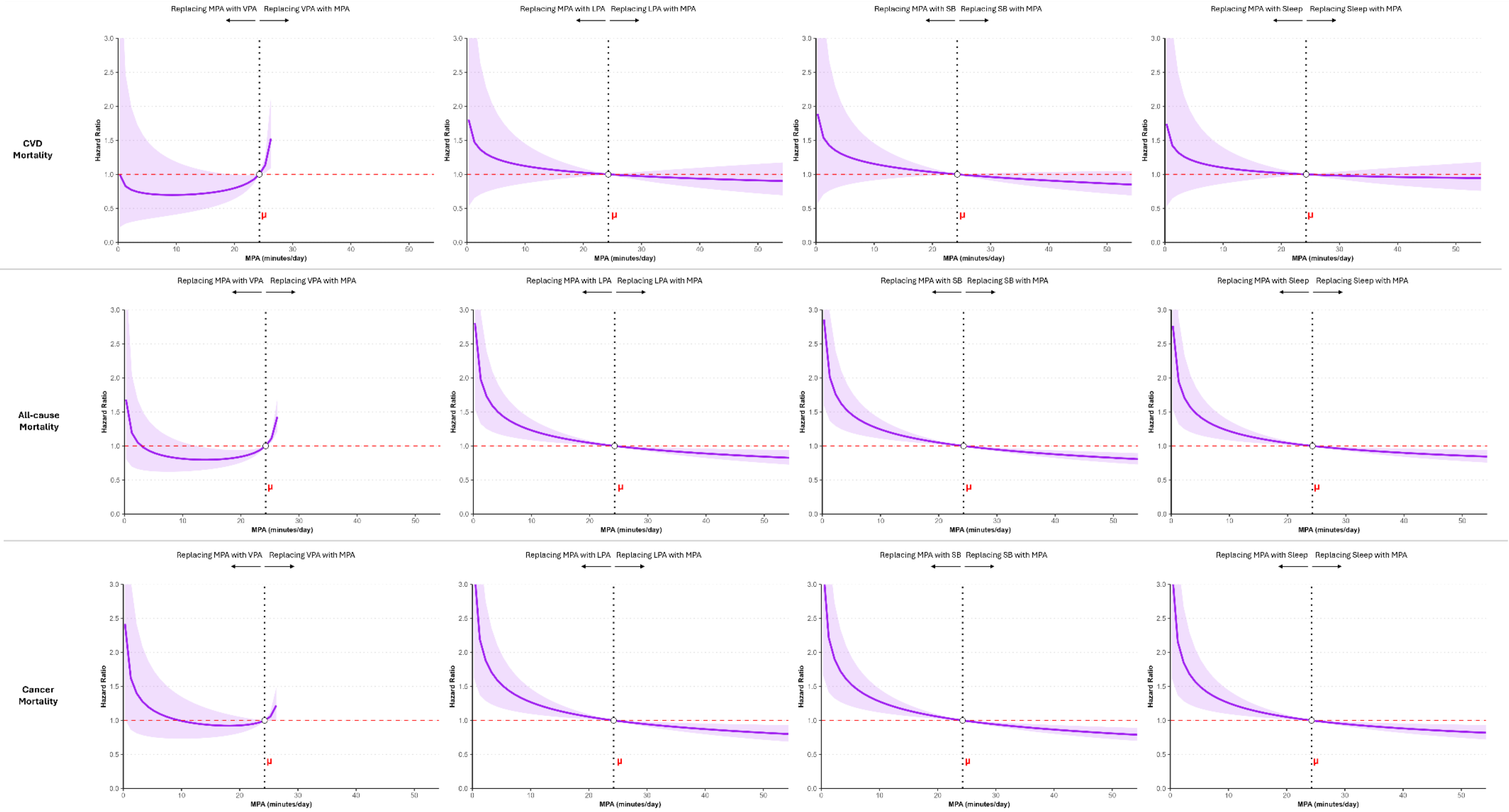

Table S19. Predicted HR of All-cause mortality amongst Cancer survivors from Isotemporal substitution around sample average of MPA whilst holding other components constant at sample average.

| | ΔRisk | MPA (daily mins) | → SB | | | MPA (daily mins) | → SLEEP | | | MPA (daily mins) | → LPA | | | | | MPA (daily mins) | → VPA | | |
|---|---|---|---|---|---|---|---|---|---|---|---|---|---|---|---|---|---|---|---|
| | | | HR | 95% CI | | | HR | 95% CI | | | HR | 95% CI | | | | | HR | 95% CI | |
| Decreasing MPA | **25%** | 10 | **1.24** | 1.11 | 1.38 | 9 | **1.24** | 1.09 | 1.41 | 9 | **1.25** | 1.09 | 1.43 | Decreasing MPA | **-25%** | - | **-** | - | - |
| | **20%** | 11 | **1.21** | 1.09 | 1.33 | 10 | **1.21** | 1.08 | 1.36 | 11 | **1.20** | 1.07 | 1.33 | | **-20%** | 15 | **0.80** | 0.67 | 0.96 |
| | **15%** | 14 | **1.14** | 1.07 | 1.22 | 13 | **1.14** | 1.05 | 1.24 | 13 | **1.15** | 1.06 | 1.25 | | **-15%** | 20 | **0.85** | 0.76 | 0.94 |
| | **10%** | 16 | **1.10** | 1.05 | 1.16 | 15 | **1.11** | 1.04 | 1.18 | 16 | **1.10** | 1.04 | 1.16 | | **-10%** | 22 | **0.90** | 0.84 | 0.95 |
| | **5%** | 20 | **1.05** | 1.02 | 1.07 | 19 | **1.05** | 1.02 | 1.08 | 19 | **1.06** | 1.02 | 1.09 | | **-5%** | 23 | **0.94** | 0.91 | 0.97 |
| Increasing MPA | **-5%** | 29 | **0.95** | 0.93 | 0.98 | 31 | **0.95** | 0.92 | 0.98 | 30 | **0.95** | 0.92 | 0.98 | Increasing MPA | **5%** | - | **-** | - | - |
| | **-10%** | 36 | **0.90** | 0.86 | 0.95 | 39 | **0.90** | 0.85 | 0.96 | 38 | **0.90** | 0.84 | 0.96 | | **10%** | 25 | **1.11** | 1.05 | 1.17 |
| | **-15%** | 45 | **0.85** | 0.78 | 0.92 | 52 | **0.85** | 0.77 | 0.94 | 48 | **0.85** | 0.76 | 0.95 | | **15%** | - | **-** | - | - |
| | **-20%** | 56 | **0.80** | 0.72 | 0.89 | 71 | **0.80** | 0.69 | 0.93 | 62 | **0.80** | 0.69 | 0.93 | | **20%** | - | **-** | - | - |
| | **-25%** | 69 | **0.75** | 0.66 | 0.86 | 102 | **0.75** | 0.61 | 0.92 | 81 | **0.75** | 0.61 | 0.92 | | **25%** | - | **-** | - | - |

Table S20. Predicted HR of Subsequent Cancer mortality amongst Cancer survivors from Isotemporal substitution around sample average of MPA whilst holding other components constant at sample average.

| | | MPA | → SB | | | MPA | → SLEEP | | | MPA | → LPA | | | | | MPA | → VPA | | |
|---|---|---|---|---|---|---|---|---|---|---|---|---|---|---|---|---|---|---|---|
| | ΔRisk | (daily mins) | HR | 95% CI | | (daily mins) | HR | 95% CI | | (daily mins) | HR | 95% CI | | | | (daily mins) | HR | 95% CI | |
| **Decreasing MPA** | **25%** | 11 | **1.24** | 1.10 | 1.39 | 10 | **1.25** | 1.09 | 1.43 | 10 | **1.26** | 1.09 | 1.45 | **Decreasing MPA** | **-25%** | - | **-** | - | - |
| | **20%** | 12 | **1.21** | 1.09 | 1.34 | 12 | **1.19** | 1.07 | 1.32 | 12 | **1.20** | 1.07 | 1.35 | | **-20%** | - | **-** | - | - |
| | **15%** | 14 | **1.16** | 1.07 | 1.26 | 14 | **1.15** | 1.05 | 1.24 | 14 | **1.15** | 1.06 | 1.26 | | **-15%** | - | **-** | - | - |
| | **10%** | 17 | **1.10** | 1.04 | 1.16 | 16 | **1.11** | 1.04 | 1.18 | 17 | **1.10** | 1.04 | 1.16 | | **-10%** | - | **-** | - | - |
| | **5%** | 20 | **1.05** | 1.02 | 1.08 | 20 | **1.05** | 1.02 | 1.08 | 20 | **1.05** | 1.02 | 1.08 | | **-5%** | - | **-** | - | - |
| **Increasing MPA** | **-5%** | 29 | **0.95** | 0.92 | 0.98 | 29 | **0.95** | 0.93 | 0.98 | 29 | **0.95** | 0.92 | 0.98 | **Increasing MPA** | **5%** | - | **-** | - | - |
| | **-10%** | 35 | **0.90** | 0.85 | 0.95 | 37 | **0.90** | 0.84 | 0.96 | 35 | **0.90** | 0.84 | 0.96 | | **10%** | - | **-** | - | - |
| | **-15%** | 42 | **0.85** | 0.78 | 0.93 | 46 | **0.85** | 0.77 | 0.94 | 44 | **0.85** | 0.76 | 0.95 | | **15%** | - | **-** | - | - |
| | **-20%** | 52 | **0.80** | 0.71 | 0.90 | 60 | **0.80** | 0.69 | 0.92 | 54 | **0.80** | 0.69 | 0.93 | | **20%** | 25 | **1.22** | 1.00 | 1.48 |
| | **-25%** | 64 | **0.75** | 0.65 | 0.87 | 79 | **0.75** | 0.62 | 0.91 | 68 | **0.75** | 0.61 | 0.92 | | **25%** | - | **-** | - | - |

Table S21. Association of Coordinates of daily-movement behaviours (intensities, including MVPA) and sleep with risk of CVD-mortality among previous cancer survivors (Cox Proportional Hazards Model) excluding all self-reported history of cancer (n=2185), any cancer events within 2y prior to accelerometry measurement (n=481), and events within 2y of follow-up (n=27).

| | Age and Sex-adjusted | | | | Fully adjusted | | | |
|---|---|---|---|---|---|---|---|---|
| | HR | 95% CI | | p-value | HR | 95% CI | | p-value |
| **Moderate-to-Vigorous PA** (vs all other behaviours) | **0.77** | **0.66** | **0.90** | **0.001** | **0.79** | **0.67** | **0.95** | **0.009** |
| Light PA (vs all other behaviours) | 0.71 | 0.38 | 1.31 | 0.271 | 0.74 | 0.40 | 1.40 | 0.361 |
| Sleep (vs all other behaviours) | 0.62 | 0.37 | 1.06 | 0.080 | 0.62 | 0.36 | 1.07 | 0.086 |
| **Sedentary behaviour** (vs all other behaviours) | **2.93** | **1.61** | **5.34** | **0.009** | **2.71** | **1.46** | **5.02** | **0.002** |

**Coefficients are ILR transformed units and not directly interpretable.**

Table S22. Association of Coordinates of daily-movement behaviours (intensities, including MVPA) and sleep with risk of All-cause mortality among previous cancer survivors (Cox Proportional Hazards Model) excluding all self-reported history of cancer (n=2185), all cancer events within 2y prior to accelerometry measurement (n=481), and events within 2y follow-up (n=27).

| | Age and Sex-adjusted | | | | Fully adjusted | | | |
|---|---|---|---|---|---|---|---|---|
| | HR | 95% CI | | p-value | HR | 95% CI | | p-value |
| **Moderate-to-Vigorous PA** (vs all other behaviours) | **0.80** | **0.73** | **0.87** | **<0.001** | **0.82** | **0.75** | **0.89** | **<0.001** |
| Light PA (vs all other behaviours) | 0.74 | 0.54 | 1.02 | 0.062 | 1.00 | 0.56 | 1.05 | 0.102 |
| Sleep (vs all other behaviours) | 1.10 | 0.80 | 1.50 | 0.560 | 1.10 | 0.81 | 1.51 | 0.532 |
| **Sedentary behaviour** (vs all other behaviours) | **1.54** | **1.11** | **2.13** | **0.010** | **1.44** | **1.04** | **2.00** | **0.027** |

**Coefficients are ILR transformed units and not directly interpretable.**

Table S23. Association of Coordinates of daily-movement behaviours (intensities, including MVPA) and sleep with risk of cancer-mortality among previous cancer survivors (Cox Proportional Hazards Model) excluding all self-reported history of cancer (n=2185), all cancer events within 2y prior to accelerometry measurement (n=481), and events within 2y follow-up (n=27).

| | **Age and Sex-adjusted** | | | | **Fully adjusted** | | | |
|---|---|---|---|---|---|---|---|---|
| | **HR** | **95% CI** | | **p-value** | **HR** | **95% CI** | | **p-value** |
| **Moderate-to-Vigorous PA** (vs all other behaviours) | **0.82** | **0.74** | **0.92** | **<0.001** | **0.85** | **0.77** | **0.95** | **0.004** |
| Light PA (vs all other behaviours) | 0.76 | 0.53 | 1.12 | 0.158 | 0.79 | 0.54 | 1.15 | 0.215 |
| Sleep (vs all other behaviours) | 1.13 | 0.78 | 1.65 | 0.513 | 1.12 | 0.77 | 1.63 | 0.552 |
| Sedentary behaviour (vs all other behaviours) | 1.40 | 0.95 | 2.06 | 0.088 | 1.33 | 0.90 | 1.96 | 0.146 |

**Coefficients are ILR transformed units and not directly interpretable.**

Tab**le S24**. Association of Coordinates of daily-movement behaviours (intensities, including MVPA) and sleep with risk of CVD-mortality among previous cancer survivors (Cox Proportional Hazards Model) excluding all self-reported history of cancer (n=2185) and all non-melanoma skin cancers (n=674)

| | **Age and Sex-adjusted** | | | | **Fully adjusted** | | | |
|---|---|---|---|---|---|---|---|---|
| | **HR** | **95% CI** | | **p-value** | **HR** | **95% CI** | | **p-value** |
| **Moderate-to-Vigorous PA** (vs all other behaviours) | **0.81** | **0.70** | **0.93** | **0.004** | **0.84** | **0.73** | **0.97** | **0.002** |
| Light PA (vs all other behaviours) | 0.83 | 0.49 | 1.41 | 0.488 | 0.86 | 0.50 | 1.47 | 0.576 |
| **Sleep** (vs all other behaviours) | **0.59** | **0.39** | **0.89** | **0.012** | **0.59** | **0.39** | **0.91** | **0.018** |
| **Sedentary behaviour** (vs all other behaviours) | **2.54** | **1.49** | **4.33** | **<0.001** | **2.33** | **1.35** | **4.04** | **0.003** |

**Coefficients are ILR transformed units and not directly interpretable.**

Tabl**e S25**. Association of Coordinates of daily-movement behaviours (intensities, including MVPA) and sleep with risk of Cancer-mortality among previous cancer survivors (Cox Proportional Hazards Model) excluding all self-reported history of cancer (n=2185) and all non-melanoma skin cancers (n=674)

| | **Age and Sex-adjusted** | | | | **Fully adjusted** | | | |
|---|---|---|---|---|---|---|---|---|
| | **HR** | **95% CI** | | **p-value** | **HR** | **95% CI** | | **p-value** |
| **Moderate-to-Vigorous PA** (vs all other behaviours) | **0.80** | **0.74** | **0.86** | **<0.001** | **0.81** | **0.76** | **0.88** | **<0.001** |
| Light PA (vs all other behaviours) | 0.83 | 0.72 | 1.21 | 0.596 | 0.86 | 0.72 | 1.22 | 0.641 |
| Sleep (vs all other behaviours) | 0.98 | 0.77 | 1.26 | 0.894 | 0.99 | 0.77 | 1.27 | 0.940 |
| **Sedentary behaviour** (vs all other behaviours) | **1.37** | **1.04** | **1.80** | **0.024** | **1.32** | **1.00** | **1.74** | **0.048** |

**Coefficients are ILR transformed units and not directly interpretable.**

Tabl**e S26**. Association of Coordinates of daily-movement behaviours (intensities, including MVPA) and sleep with risk of All-cause mortality among previous cancer survivors (Cox Proportional Hazards Model) excluding all self-reported history of cancer (n=2185) and all non-melanoma skin cancers (n=674)

| | Age and Sex-adjusted | | | | Fully adjusted | | | |
|---|---|---|---|---|---|---|---|---|
| | **HR** | **95% CI** | | **p-value** | **HR** | **95% CI** | | **p-value** |
| **Moderate-to-Vigorous PA** (vs all other behaviours) | **0.79** | **0.74** | **0.84** | **<0.001** | **0.80** | **0.75** | **0.85** | **<0.001** |
| Light PA (vs all other behaviours) | 0.98 | 0.78 | 1.11 | 0.897 | 1.00 | 0.79 | 1.27 | 0.998 |
| Sleep (vs all other behaviours) | 0.90 | 0.72 | 1.12 | 0.347 | 0.80 | 0.73 | 1.13 | 0.406 |
| **Sedentary behaviour** (vs all other behaviours) | **1.44** | **1.13** | **1.83** | **0.004** | **1.38** | **1.08** | **1.76** | **0.011** |

**Coefficients are ILR transformed units and not directly interpretable.**

| Table S27. E-Values (estimate of minimal association of a residual confounder with the exposure and outcome (All-cause mortality), required to nullify the associations observed in the fully-adjusted model). | | | |
|---|---|---|---|
| **Exposure** | E-Value | Lower CI | Upper CI |
| **Moderate-to-vigorous physical activity** (vs all other postures) | 1.83 | - | 1.67 |
| **Sedentary Behaviour** (vs all other postures) | 2.11 | 1.497 | - |

| Table S28. E-Values (estimate of minimal association of a residual confounder with the exposure and outcome (Cancer-mortality), required to nullify the associations observed in the fully-adjusted model). | | | |
|---|---|---|---|
| **Exposure** | E-Value | Lower CI | Upper CI |
| **Moderate-to-vigorous physical activity** (vs all other postures) | 1.76 | - | 1.56 |
| **Sedentary Behaviour** (vs all other postures) | 1.91 | 1.16 | - |

Table S29. E-Values (estimate of minimal association of a residual confounder with the exposure and outcome (CVD-mortality), required to nullify the associations observed in the fully-adjusted model).

| **Exposure** | E-Value | Lower CI | Upper CI |
|---|---|---|---|
| **Moderate-to-vigorous physical activity** (vs all other postures) | 1.73 | - | 1.39 |
| **Sleep duration** (vs all other postures) | 2.45 | - | 1.38 |
| **Sedentary Behaviour** (vs all other postures) | 3.14 | 1.75 | - |
